# Development and validation of a multivariate model for predicting heart failure hospitalization and mortality in patients receiving maintenance hemodialysis

**DOI:** 10.1101/2023.03.14.23287278

**Authors:** Wenwu Tang, Xinzhu Yuan, Zhixin Wang, Ying Zhang, Xiaoxia Chen, Xiaohua Yang, Zhirui Qi, Ju Zhang, Jie Li, Xisheng Xie

**Author notes:** W.Tang 、X.Yuan and Z.Wang contributed equally. Correspondence: Xisheng Xie,.

## Abstract

**Background:** Heart failure (HF) is a common cardiovascular disease in patients receiving maintenance hemodialysis (MHD). Once these patients on MHD exhibit HF, their hospitalization rate, mortality, and economic burden will be significantly increased. Early identification and prediction of hospitalization and death are of great importance for reducing hospitalization and mortality. This study used multicenter clinical data to develop and externally validate clinical risk models to predict expected mortality and HF hospitalization rates in MHD patients with HF.

**Materials and Methods:** From January 2017 to October 2022, 348 patients receiving MHD from four participating centers were enrolled. Demographic data, MHD treatment modalities, laboratory tests, and echocardiography data were collected when the initial event occurred. Three centers were randomly assigned to the modeling dataset (n=258), and one center was assigned to the external validation set (n=90). Considering a composite outcome of HF hospitalization and death as the primary endpoint and hospitalization due to HF or death as the secondary endpoint, a COX clinical prediction model was constructed and verified using internal and external datasets.

**Results:** The median age of patients in the modeling cohort was 63 years old, 41.5% of patients were women; 165 (61%) had a history of HF; 81 (31.4%) were hospitalized for HF; and 39 (15.1%) patients had died. The c-statistic values for composite outcome, hospitalization for HF, and mortality were 0.812, 0.808, and 0.811, respectively. The predictors of death and hospitalization outcomes caused by HF are significantly different. The strongest predictors of HF hospitalization outcomes were advanced age, multiple HF hospitalizations, hyponatremia, high levels of NT-proBNP and hs-cTnT, and larger MVe values. The strongest predictors of mortality were longer dialysis age, combined atrial fibrillation, calcification of the aortic or mitral valve (especially calcification, and in particular aortic valve calcification), pleural effusion, low serum sodium, and higher levels of hs-cTnT. The median age of the patients in the external validation cohort was 63 years old; 28.8% were female; 35 (38.1%) had a history of HF; 11 (12.2%) were hospitalized for HF; and 5 (5.6%) died. The c-statistic of the predictive models for composite outcome, hospitalisation for HF, and mortality was comparable to that of the modelling cohort.

**Conclusion:** The model established in this study is stable and reliable and the included variables are easily obtained from the routine clinical environment. The model can provide useful risk factors and prognostic information for patients with MHD combined with HF. Keywords: heart failure, MHD patients, mortality, predictive model, external validation.

## 1. Introduction

The number of patients receiving renal replacement therapy (RRT) worldwide exceeded 2.5 million in 2010, and is expected to increase to approximately 5.4 million by 2030^[1]^. The risk of cardiovascular disease (CVD) in patients on maintenance hemodialysis (MHD) is 20-times higher than that in the general population, and heart failure (HF) is the second most common CVD in patients undergoing MHD (about 10.2%)^[2,3]^. According to the United States Renal Data System data report, the medical insurance expenditure for patients with MHD in 2016 alone was as high as $35 billion; in addition, the 2-year mortality rate of patients with chronic kidney disease (CKD) but without HF was approximately 16.5%, while the overall 2-year mortality rate those with HF was as high as 33.3%^[4–6]^.

Compared to the general population of patients with HF, patients receiving MHD face more complex risk factors for uremic cardiomyopathy based on traditional risk factors (hypertension, diabetes) due to impaired glomerular filtration and secretory function; Furthermore, although patients receiving MHD can prolong survival with long-term RRT treatment, this also increases risks related to dialysis (such as hemodynamics, rapid changes in electrolyte composition, and micro-inflammatory state induced by dialysis materials). Therefore, it is becoming an urgent need to effectively identify individuals with HF at high-risk of hospitalization or death among these patients^[7–9]^.

Although the clinical condition is severe, it is still difficult to predict the mortality and hospitalization rate of HF patients using the published prediction models. A meta-analysis reported that only 33% of the 117 published predictive models were validated in separate cohort studies, and most studies only reported moderate predictive performance^[9,10]^. Joanne’s predictive model had a c-statistic of 0.71 and 0.70 for HF hospitalization and death, respectively^[11]^; while the c-statistics of Joshua’s prediction model were 0.80 and 0.79, respectively^[10–12]^. Unfortunately, these prediction models almost all excluded or did not focus on patients receiving MHD, so they are not suitable for this specific population. Recently, the model by Gotta et al. focused mainly on the prediction of death in patients with MHD under 30 years of age but did consider the effect of HF in these patients^[13]^. The purpose of the present study is to establish a prediction model suitable for patients with MHD to accurately predict the risk of hospitalisation and death from HF, to strengthen the early identification and intervention of clinicians, and to assist doctors and patients in making better personalized management decisions to effectively improve the prognosis of patients receiving MHD. In addition, the model also helps to select high-risk patients more accurately for future clinical trials and to enrich the reported incidence of clinical events, and reduce the sample size of future studies^[14]^.

## 2. Materials and methods

### 2.1 Materials

We used the Transparent Reporting of a Multivariate Prediction Model for Individual Prognosis or Diagnosis (TRIPOD) as a guide to develop and validate our model.

#### 2.1.1 Subjects

We enrolled subjects From January 2017 to October 2022 (with at least 60 days of follow-up), 1079 patients receiving MHD from four centers (Nanchong Central Hospital, Guangyuan Central Hospital, Suining Central Hospital and Peng’an County People’s Hospital). Patients with CKD5 dialysis who met the Kidney Disease Improving Global Outcomes (KDIGO) criteria developed by the American Society of Nephrology, and regular hemodialysis time ≥3 months, at least 2 times a week, and met the clinical diagnostic criteria determined by the International RLS Study Group in 2014 were enrolled.

#### 2.1.2 Inclusion criteria

All patients (1) aged >18 years old; (2) who had received MHD treatment for more than 3 months; (3) conformed to the diagnostic criteria of HF; (4) who had provided informed consent and voluntary participation in this study; and (5) those with complete clinical data were selected for the final analysis.

#### 2.1.3 Exclusion criteria

Patients meeting the following criteria were excluded: (i) having a history of liver disease, malignant tumor, mental illness, or other serious diseases; (ii) patients who were unable to cooperate, unwilling to participate, or had incomplete clinical data. Following the application of the inclusion and exclusion criteria, 693 patients without hospitalization history and 38 patients with missing biochemical or echocardiographic data were excluded, and 348 patients were included in the study (Figure 1).

**Figure 1.**
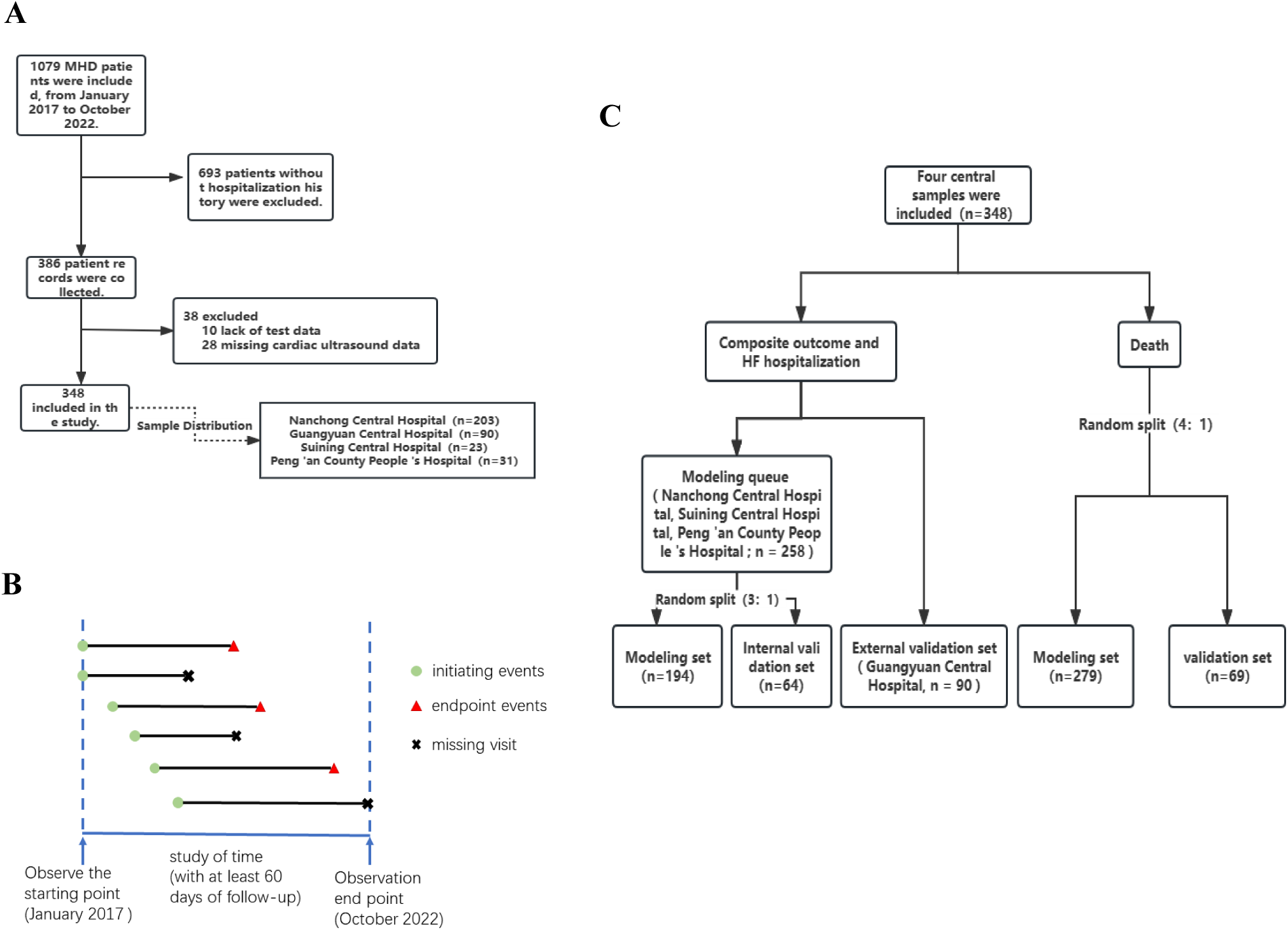
Research flow chart. (A) Patient inclusion process according to nadir criteria; (B) schematic diagram of start-and endpoint events; (C) schematic diagram of allocation of the modelling and validation datasets.

### 2.2 Methods

#### 2.1.1 Diagnostic criteria for HF

The following criteria were used to diagnose HF in this study: (a) new or worsening HF symptoms or previous hospitalisation history; (b) left ventricular ejection fraction (LVEF) ≤40%; if LVEF >40%, abnormal N-terminal B-type natriuretic peptide (NT-proBNP) (>11215.2 pg/mL), (c) with either ‘left ventricular hypertrophy and/or left atrial enlargement’ or ‘cardiac diastolic dysfunction’. A diagnosis of HF was made by meeting either a/b or a/c of the above criteria. The diagnosis of HF was made by two trained physicians, and a specialist was consulted in case of disagreement.

#### 2.1.2 NT-proBNP

In this study, NT-proBNP > 11215.2 pg/mL was selected as the diagnostic threshold for HF in MHD patients.

#### 2.2.3 Starting point and endpoint of observation

The time of first hospitalization of the patient was taken as the starting point of observation, and the times of hospitalization due to HF, death, transfer, renal transplantation, and arrival after follow-up was taken as the endpoint of observation. The time between the starting point and the endpoint was considered as the follow-up time. In this study, the composite outcome of HF hospitalization and death was the primary endpoint, and HF hospitalization or death was the secondary endpoint (Figure 1).

#### 2.2.4 Study Indicators

A total of 112 variables were included in this study. (1) General and dialysis-related data: including sex, age, dialysis age, type of dialysis vascular access, reasons for dialysis (diabetic nephropathy, glomerular inflammatory disease, or hypertension), previous cumulative HF hospitalization times and total hospitalization days, body mass index (BMI), dialysis times (weekly), dialysis duration (h/time). (2) Clinical data: vital signs at admission (pulse, pulse pressure difference), typical symptoms at HF (exertional or nocturnal paroxysmal dyspnea, sitting breathing, edema, rales, third heart sound, jugular vein filling, or positive hepatic neck reflux sign), history of coronary heart disease or coronary stent surgery, other surgical history (thyroid radiofrequency ablation or arterial dissection), history of cerebrovascular accident (cerebral infarction or cerebral hemorrhage), history of atrial fibrillation, bradycardia (conduction block), metabolic acidosis, infection (pneumonia, urinary tract infection, dialysis catheter-related infection, and sepsis), thyroid function, history of diabetes and diabetes-related complications (such as diabetic retinopathy, and diabetic peripheral neuropathy), chronic obstructive pulmonary disease (COPD), history of other lung diseases (respiratory failure or pulmonary edema, bronchitis, bronchiectasis, emphysema or bullae, silicosis, pneumoconiosis, lung damage, pulmonary fibrosis), liver or pancreatic diseases (fatty liver, cirrhosis and decompensation, pancreatic injury, hepatitis B), history of cholecystitis or cholecystectomy, gastrointestinal ulcer, gastrointestinal inflammation and diarrhea, gastrointestinal bleeding, history of serous cavity effusion (pericardium, chest, pelvis), coagulation abnormalities or less platelets, immunodeficiency, tumor, NYHA classification, HF stage, and length of hospital stay at the beginning of the event. (3) Laboratory examination: white blood cell count, C-reactive protein, hemoglobin, parathyroid hormone, NT-proBNP, high sensitivity troponin T (hs-cTnT), total protein, albumin, uric acid, creatinine, cystatin, triglyceride, total cholesterol, high density lipoprotein cholesterol, low density lipoprotein cholesterol, aspartate aminotransferase (AST), alanine aminotransferase (ALT), total bilirubin, serum potassium, serum sodium, serum calcium, and serum phosphorus. (4) Echocardiographic data: left atrial changes, left ventricular enlargement (atrial and ventricular), right atrial changes, right ventricular enlargement, whole heart enlargement, aortic or pulmonary artery changes (widened), interventricular septum changes (thickened), left ventricular wall changes (hypertrophy), ventricular wall motion ability (reverse movement of interventricular septum and left ventricular posterior wall, and diffuse weakening of left ventricular wall), calcification of aortic valve or mitral valve, valvular regurgitation (aortic valve, pulmonary valve, mitral valve, and tricuspid valve), left ventricular diastolic dysfunction, left ventricular or right ventricular systolic dysfunction, pulmonary hypertension, pericardial effusion (mm), left atrium (LA), left ventricle (LV), right atrium (RA), right ventricle (RV), interventricular septum (IVS), left ventricular posterior wall (LVPW), main pulmonary artery (MPA), aorta or aorta root (AO), ascending aorta (AAO), anterior mitral flow velocity (MVe), aortic valve (AV), fractional shortening (FS), and LVEF. The reasons for including the above variables are as follows: First, factors related to death and hospitalization rates have been identified in combination with guidelines and related studies. Secondly, compared to non-MHD patients, risk factors for HF in MHD patients were more complex. This study collected data relative to the above variables based on the data availability of each center and cases with missing data were excluded.

### 2.3. Design of the prediction model

#### 2.3.1 Construction and evaluation of prediction model

Patients were divided into training and test groups based on center enrolment, and the clinical features were selected from all independent variables. The importance of each index in different model training and test groups was compared using a COX prediction model, and the best model was then used for evaluation and verification. The detailed steps are described below.

##### 2.3.1.1 Allocation of modeling and validation sets

The four centers were divided into two groups with the composite endpoint of “death and HF hospitalization” or “HF hospitalization”. Three centers were assigned to the modelling set (n=258) and one center was assigned to the validation set (n=90). The patients in the modelling cohort (n=258) were then divided into a modelling set (n=194) and an internal validation set (n=64) in a 3:1 ratio. The median follow-up for the modeling cohort was 10 months (IQR, 5– 16 months). Because the sample size of death events included only 44 cases, for the model established with “death” as the outcome, all patients (n=348) were randomly divided into the modeling cohort (n=279) and the internal validation cohort (n=69) according to the ratio of 4:1. The median follow-up time of the modeling cohort was 12 months (IQR: 7–20 months). The allocation process is detailed in Figure 1.

##### 2.3.1.2 Statistical analysis of clinical features for screening patients

SPSS v26.0 (IBM, USA)was used for COX univariate analysis and R software (University of Auckland, New Zealand, glmnet 4.2.2) for Least Absolute Shrinkage and Selection Operator (LASSO) regression analysis. LASSO regression can compress variable coefficients, prevent overfitting, and resolve serious collinearity concerns^[15]^. SPSS v26.0 was used for multivariate COX regression analysis, and finally the characteristic variables with two-tailed P-values <0.05 were obtained ^[16]^.

##### 2.3.1.3 Model verification

R software (version 4.2.2) and the “survivalROC” package were used to construct receiver operating characteristic (ROC) curves of the modeling set and the validation set, and the area under the curve and c-statistics were calculated to validate the accuracy of the prediction model ^[17]^. The R software package “survivalROC” was used to draw calibration curves to measure the predictive ability of the model and to reflect the concordance between the predicted risk and the actual risk^[17]^. To assess the clinical applicability of the prediction model, the R software package ‘stdca’ was used to perform decision curve analysis (DCA)^[18]^.

### 2.4 Statistical analysis

Variables included in the modelling and validation sets were compared. Continuous variables were expressed as median and interquartile ranges (IQR) and compared using the Mann–Whitney U test. Categorical variables were expressed as numbers and percentages and compared using the chi-square test. A statistically significant differences was defined by two-sided tests with a P-value<0.05. SPSS v26.0 and the R software package v4.2.2 were used for statistical analysis.

## 3. Results

### 3.1 Comparison of baseline data

All variables were investigated for all patients at the time of the initiation event: laboratory tests and ultrasound data were the first results after admission, and data related to dialysis treatment were the closest to the pre-initiation schedule. Hyponatremia was defined as a serum sodium level of 135 mmol/L. Infections included pneumonia, sepsis, urinary tract, or dialysis catheter-related infections. The specific baseline data for the modelling set and the external validation set are shown in Table 1. The differences between the two sets were not statistically significant (P >0.05).

**Table 1.**
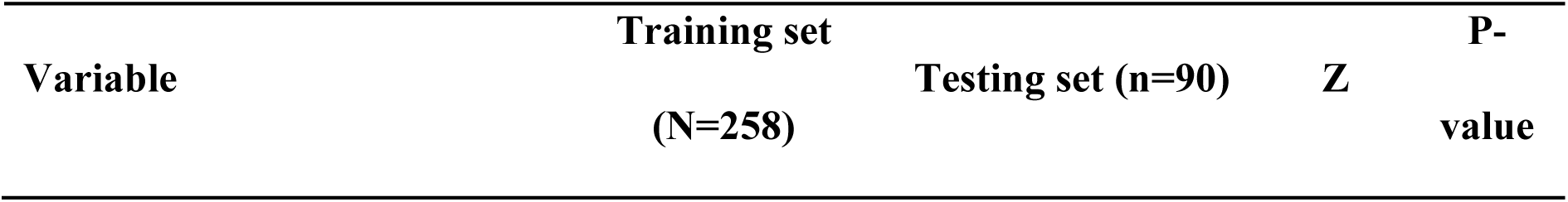
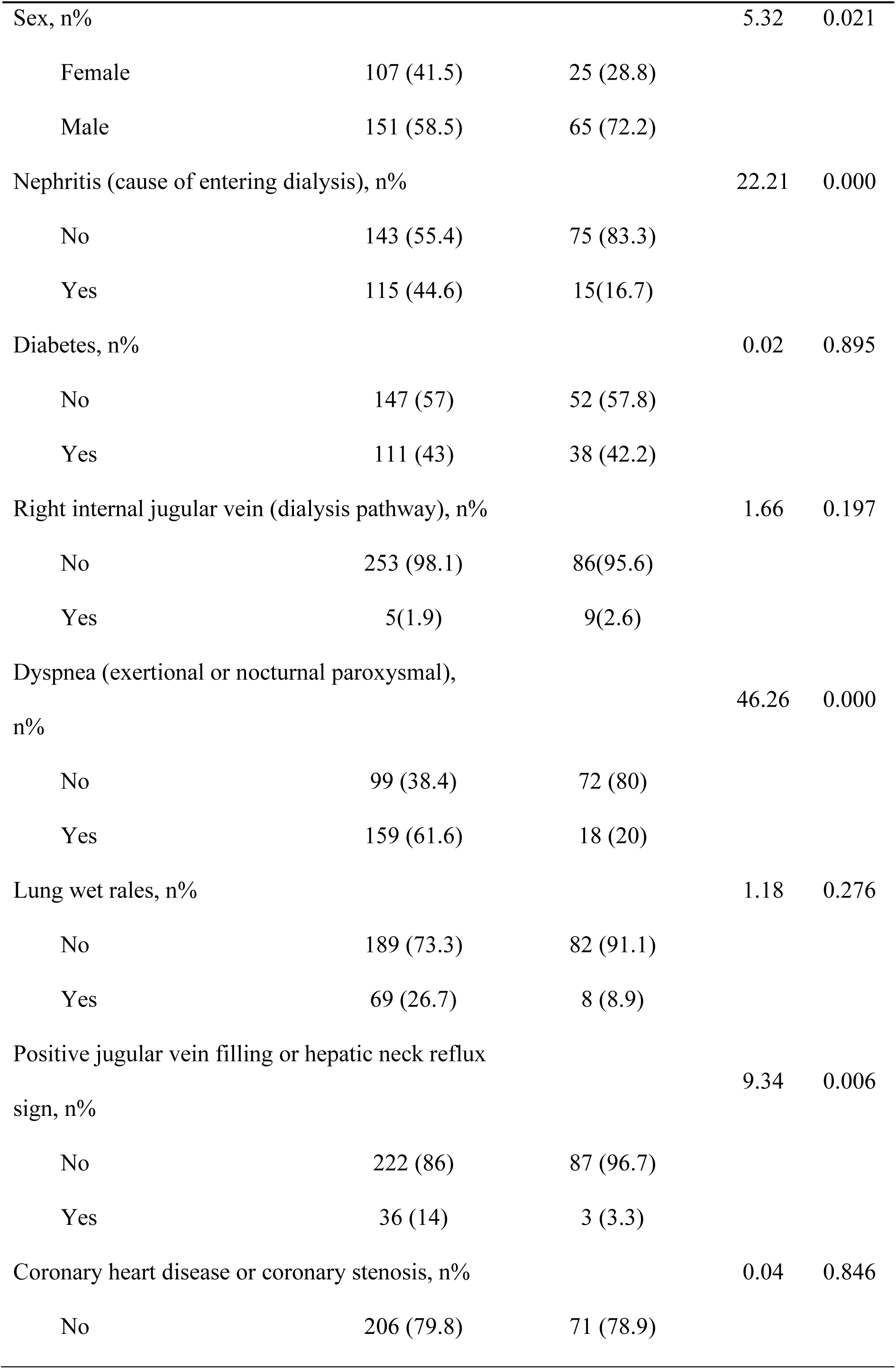
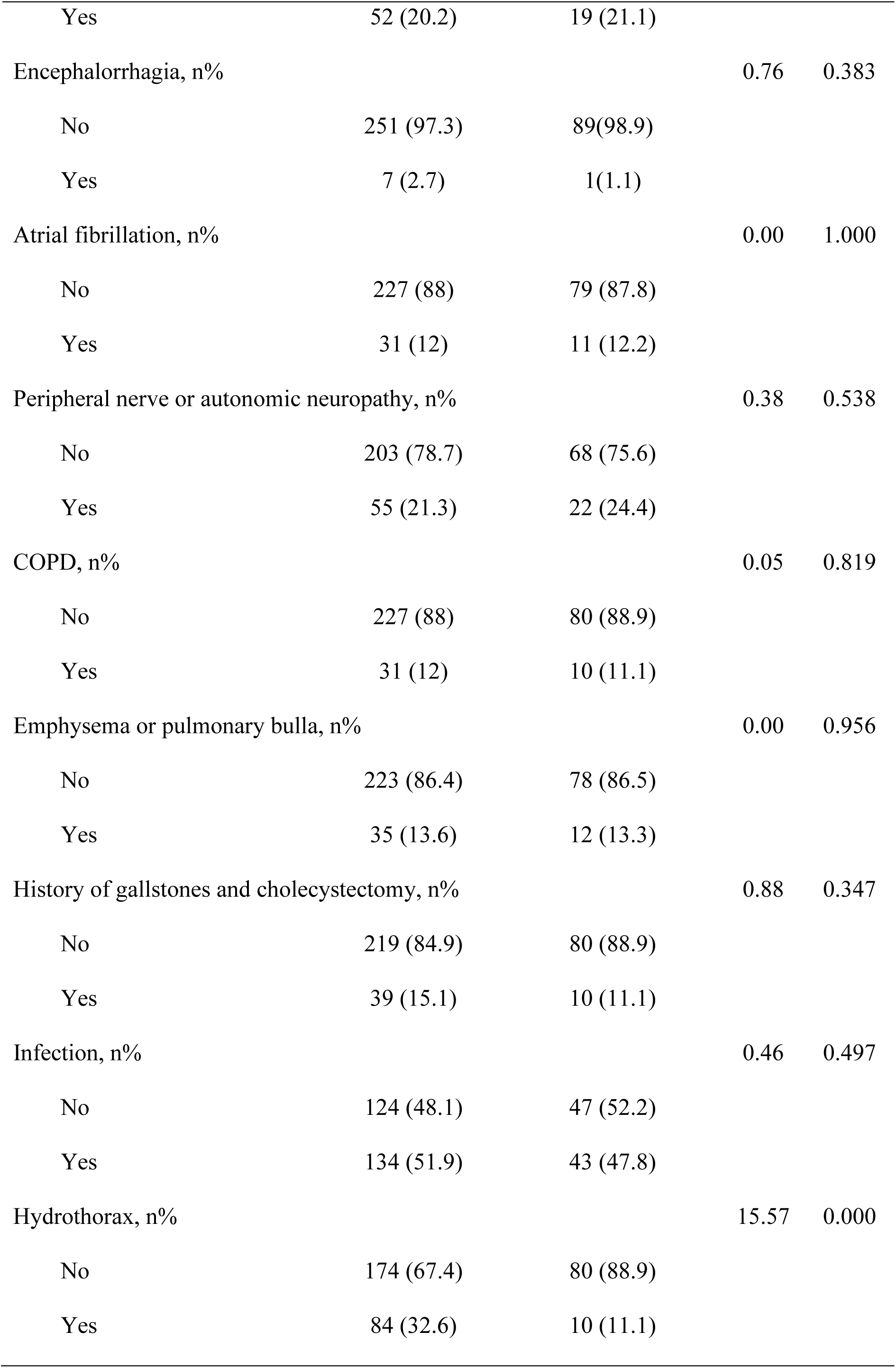
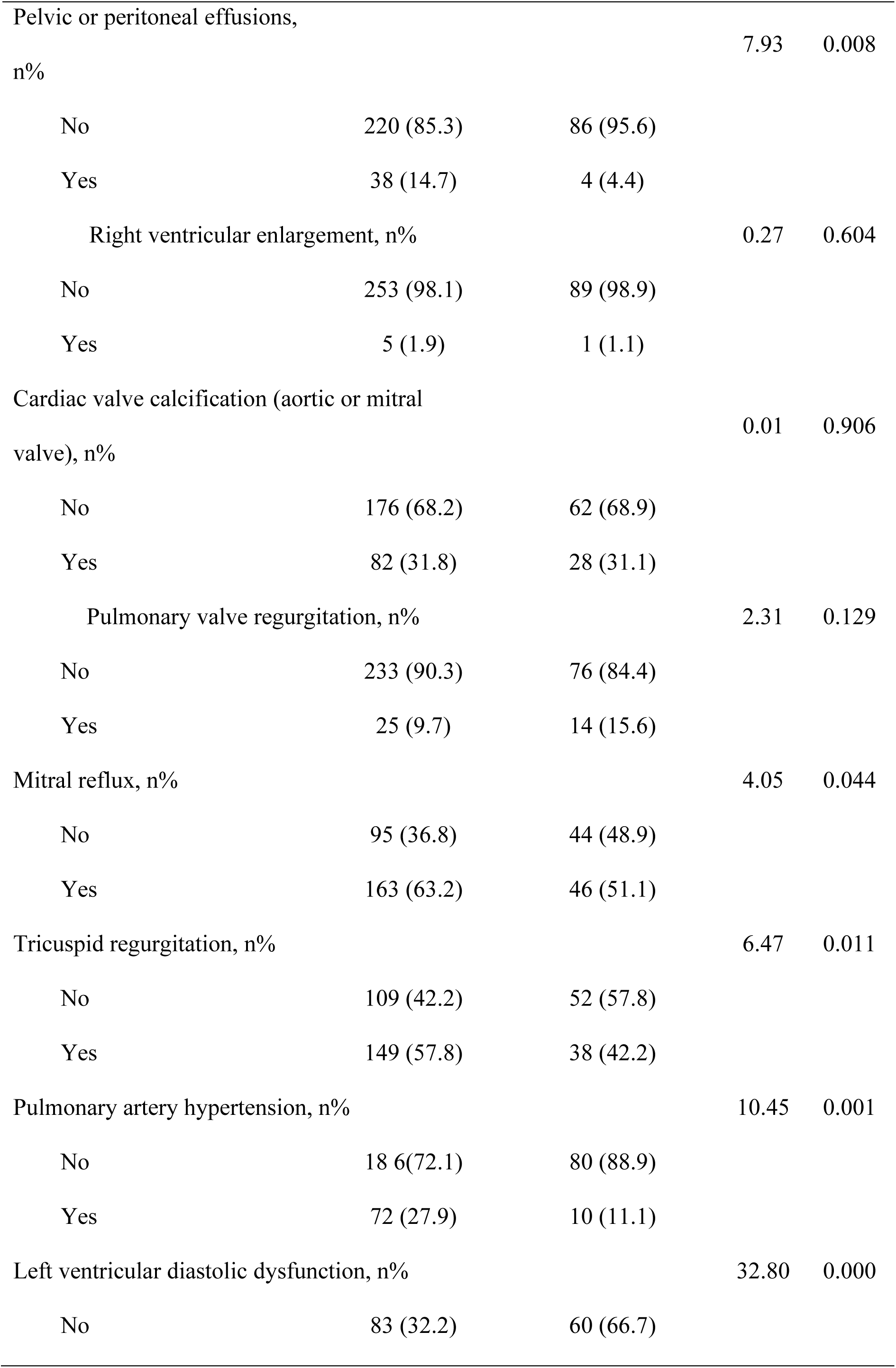
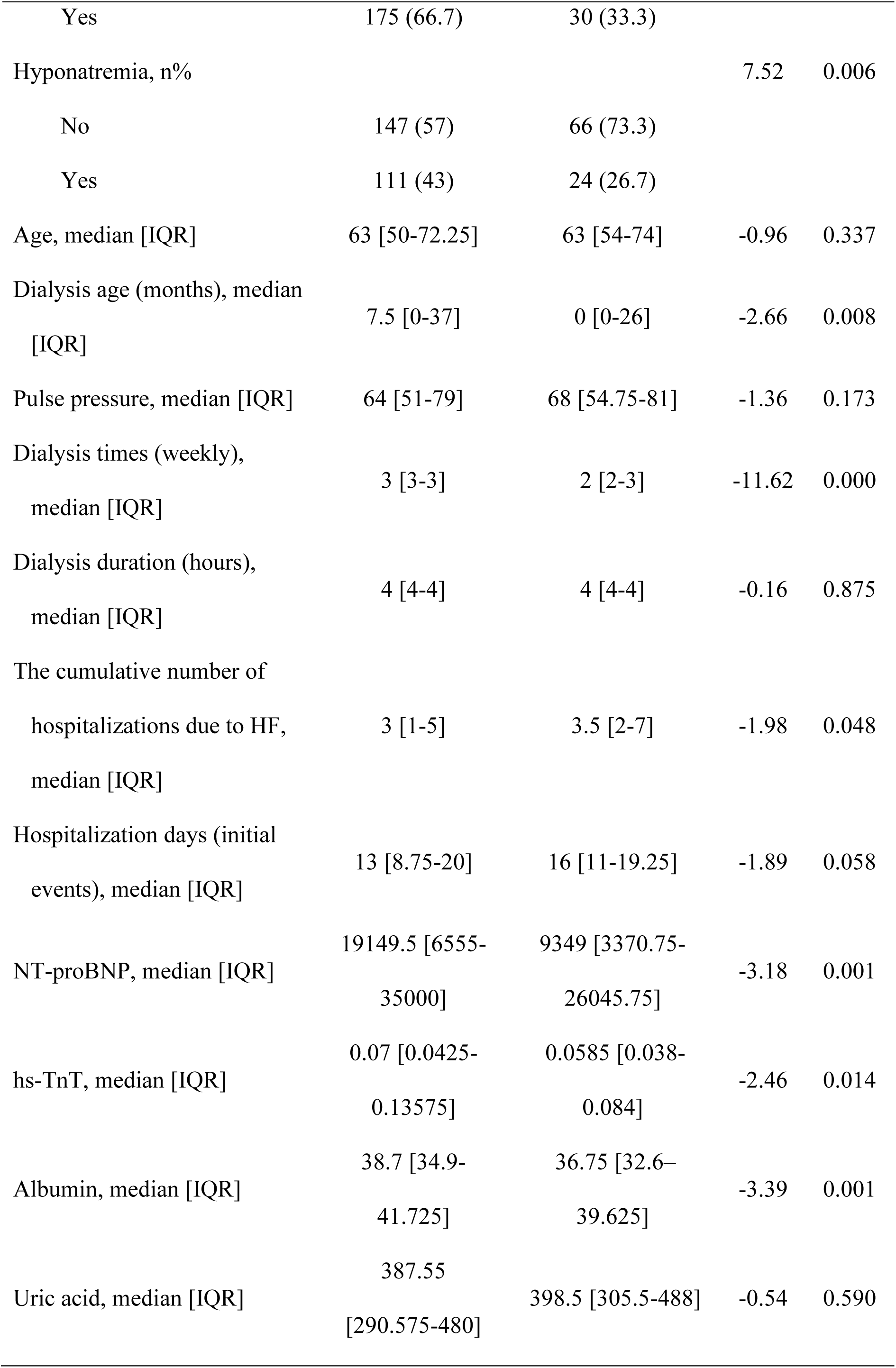
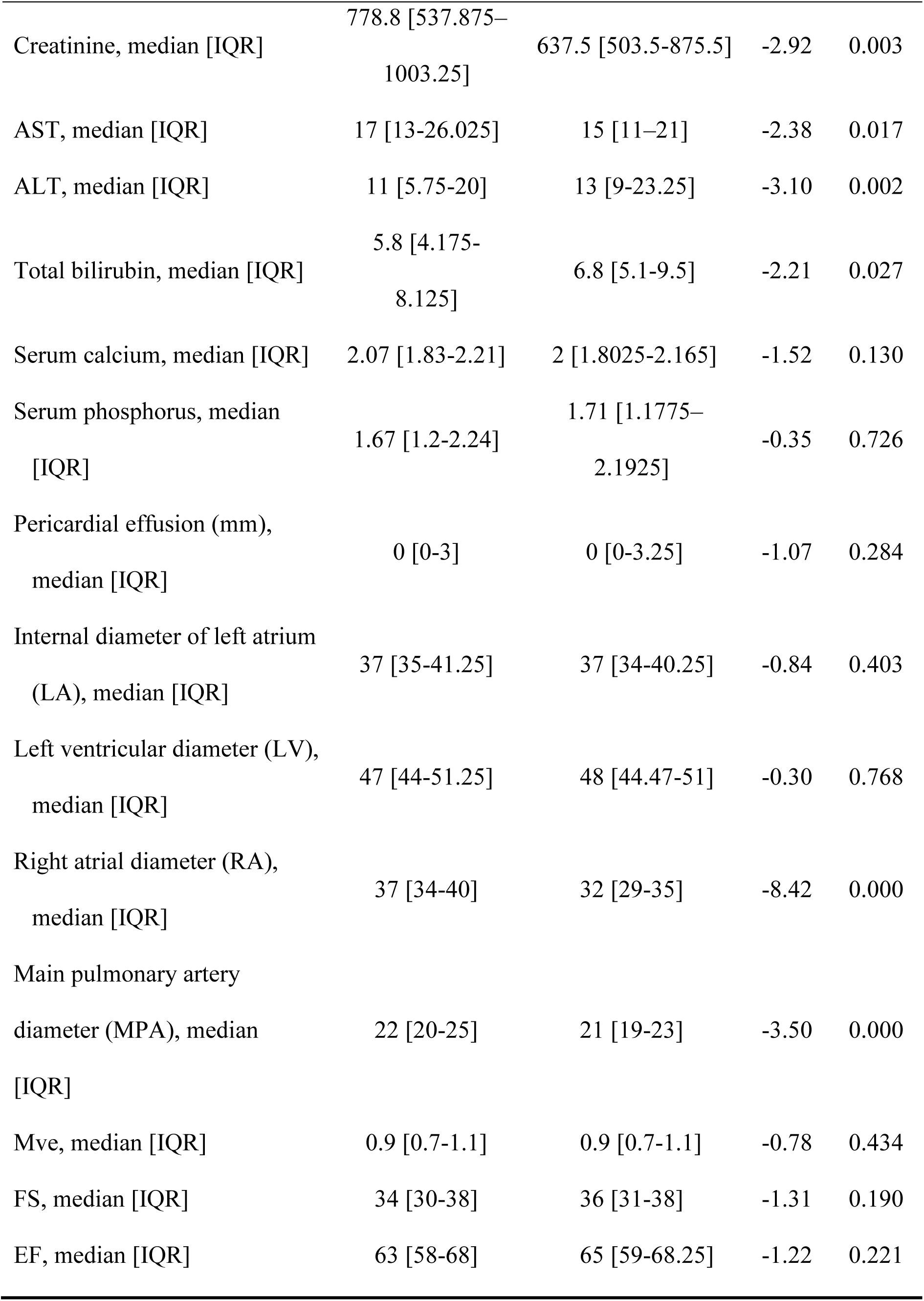
Baseline characteristics of the modeling and external validation sets

| Variable | Training set<br>(N=258) | Testing set (n=90) | Z | P-value |
| --- | --- | --- | --- | --- |
| Sex, n% |  |  | 5.32 | 0.021 |
| Female | 107 (41.5) | 25 (28.8) |  |  |
| Male | 151 (58.5) | 65 (72.2) |  |  |
| Nephritis (cause of entering dialysis), n% |  |  | 22.21 | 0.000 |
| No | 143 (55.4) | 75 (83.3) |  |  |
| Yes | 115 (44.6) | 15(16.7) |  |  |
| Diabetes, n% |  |  | 0.02 | 0.895 |
| No | 147 (57) | 52 (57.8) |  |  |
| Yes | 111 (43) | 38 (42.2) |  |  |
| Right internal jugular vein (dialysis pathway), n% |  |  | 1.66 | 0.197 |
| No | 253 (98.1) | 86(95.6) |  |  |
| Yes | 5(1.9) | 9(2.6) |  |  |
| Dyspnea (exertional or nocturnal paroxysmal),<br>n% |  |  | 46.26 | 0.000 |
| No | 99 (38.4) | 72 (80) |  |  |
| Yes | 159 (61.6) | 18 (20) |  |  |
| Lung wet rales, n% |  |  | 1.18 | 0.276 |
| No | 189 (73.3) | 82 (91.1) |  |  |
| Yes | 69 (26.7) | 8 (8.9) |  |  |
| Positive jugular vein filling or hepatic neck reflux<br>sign, n% |  |  | 9.34 | 0.006 |
| No | 222 (86) | 87 (96.7) |  |  |
| Yes | 36 (14) | 3 (3.3) |  |  |
| Coronary heart disease or coronary stenosis, n% |  |  | 0.04 | 0.846 |
| No | 206 (79.8) | 71 (78.9) |  |  |
| Yes | 52 (20.2) | 19 (21.1) |  |  |
| Encephalorrhagia, n% |  |  | 0.76 | 0.383 |
| No | 251 (97.3) | 89(98.9) |  |  |
| Yes | 7 (2.7) | 1(1.1) |  |  |
| Atrial fibrillation, n% |  |  | 0.00 | 1.000 |
| No | 227 (88) | 79 (87.8) |  |  |
| Yes | 31 (12) | 11 (12.2) |  |  |
| Peripheral nerve or autonomic neuropathy, n% |  |  | 0.38 | 0.538 |
| No | 203 (78.7) | 68 (75.6) |  |  |
| Yes | 55 (21.3) | 22 (24.4) |  |  |
| COPD, n% |  |  | 0.05 | 0.819 |
| No | 227 (88) | 80 (88.9) |  |  |
| Yes | 31 (12) | 10 (11.1) |  |  |
| Emphysema or pulmonary bulla, n% |  |  | 0.00 | 0.956 |
| No | 223 (86.4) | 78 (86.5) |  |  |
| Yes | 35 (13.6) | 12 (13.3) |  |  |
| History of gallstones and cholecystectomy, n% |  |  | 0.88 | 0.347 |
| No | 219 (84.9) | 80 (88.9) |  |  |
| Yes | 39 (15.1) | 10 (11.1) |  |  |
| Infection, n% |  |  | 0.46 | 0.497 |
| No | 124 (48.1) | 47 (52.2) |  |  |
| Yes | 134 (51.9) | 43 (47.8) |  |  |
| Hydrothorax, n% |  |  | 15.57 | 0.000 |
| No | 174 (67.4) | 80 (88.9) |  |  |
| Yes | 84 (32.6) | 10 (11.1) |  |  |
| Pelvic or peritoneal effusions,<br>n% |  |  | 7.93 | 0.008 |
| No | 220 (85.3) | 86 (95.6) |  |  |
| Yes | 38 (14.7) | 4 (4.4) |  |  |
| Right ventricular enlargement, n% |  |  | 0.27 | 0.604 |
| No | 253 (98.1) | 89 (98.9) |  |  |
| Yes | 5 (1.9) | 1 (1.1) |  |  |
| Cardiac valve calcification (aortic or mitral<br>valve), n% |  |  | 0.01 | 0.906 |
| No | 176 (68.2) | 62 (68.9) |  |  |
| Yes | 82 (31.8) | 28 (31.1) |  |  |
| Pulmonary valve regurgitation, n% |  |  | 2.31 | 0.129 |
| No | 233 (90.3) | 76 (84.4) |  |  |
| Yes | 25 (9.7) | 14 (15.6) |  |  |
| Mitral reflux, n% |  |  | 4.05 | 0.044 |
| No | 95 (36.8) | 44 (48.9) |  |  |
| Yes | 163 (63.2) | 46 (51.1) |  |  |
| Tricuspid regurgitation, n% |  |  | 6.47 | 0.011 |
| No | 109 (42.2) | 52 (57.8) |  |  |
| Yes | 149 (57.8) | 38 (42.2) |  |  |
| Pulmonary artery hypertension, n% |  |  | 10.45 | 0.001 |
| No | 186 (72.1) | 80 (88.9) |  |  |
| Yes | 72 (27.9) | 10 (11.1) |  |  |
| Left ventricular diastolic dysfunction, n% |  |  | 32.80 | 0.000 |
| No | 83 (32.2) | 60 (66.7) |  |  |
| Yes | 175 (66.7) | 30 (33.3) |  |  |
| Hyponatremia, n% |  |  | 7.52 | 0.006 |
| No | 147 (57) | 66 (73.3) |  |  |
| Yes | 111 (43) | 24 (26.7) |  |  |
| Age, median [IQR] | 63 [50-72.25] | 63 [54-74] | -0.96 | 0.337 |
| Dialysis age (months), median [IQR] | 7.5 [0-37] | 0 [0-26] | -2.66 | 0.008 |
| Pulse pressure, median [IQR] | 64 [51-79] | 68 [54.75-81] | -1.36 | 0.173 |
| Dialysis times (weekly), median [IQR] | 3 [3-3] | 2 [2-3] | -11.62 | 0.000 |
| Dialysis duration (hours), median [IQR] | 4 [4-4] | 4 [4-4] | -0.16 | 0.875 |
| The cumulative number of hospitalizations due to HF, median [IQR] | 3 [1-5] | 3.5 [2-7] | -1.98 | 0.048 |
| Hospitalization days (initial events), median [IQR] | 13 [8.75-20] | 16 [11-19.25] | -1.89 | 0.058 |
| NT-proBNP, median [IQR] | 19149.5 [6555-35000] | 9349 [3370.75-26045.75] | -3.18 | 0.001 |
| hs-TnT, median [IQR] | 0.07 [0.0425-0.13575] | 0.0585 [0.038-0.084] | -2.46 | 0.014 |
| Albumin, median [IQR] | 38.7 [34.9-41.725] | 36.75 [32.6-39.625] | -3.39 | 0.001 |
| Uric acid, median [IQR] | 387.55 [290.575-480] | 398.5 [305.5-488] | -0.54 | 0.590 |
| Creatinine, median [IQR] | 778.8 [537.875–<br>1003.25] | 637.5 [503.5-875.5] | -2.92 | 0.003 |
| AST, median [IQR] | 17 [13-26.025] | 15 [11–21] | -2.38 | 0.017 |
| ALT, median [IQR] | 11 [5.75-20] | 13 [9-23.25] | -3.10 | 0.002 |
| Total bilirubin, median [IQR] | 5.8 [4.175-8.125] | 6.8 [5.1-9.5] | -2.21 | 0.027 |
| Serum calcium, median [IQR] | 2.07 [1.83-2.21] | 2 [1.8025-2.165] | -1.52 | 0.130 |
| Serum phosphorus, median<br>[IQR] | 1.67 [1.2-2.24] | 1.71 [1.1775–<br>2.1925] | -0.35 | 0.726 |
| Pericardial effusion (mm),<br>median [IQR] | 0 [0-3] | 0 [0-3.25] | -1.07 | 0.284 |
| Internal diameter of left atrium<br>(LA), median [IQR] | 37 [35-41.25] | 37 [34-40.25] | -0.84 | 0.403 |
| Left ventricular diameter (LV),<br>median [IQR] | 47 [44-51.25] | 48 [44.47-51] | -0.30 | 0.768 |
| Right atrial diameter (RA),<br>median [IQR] | 37 [34-40] | 32 [29-35] | -8.42 | 0.000 |
| Main pulmonary artery<br>diameter (MPA), median<br>[IQR] | 22 [20-25] | 21 [19-23] | -3.50 | 0.000 |
| Mve, median [IQR] | 0.9 [0.7-1.1] | 0.9 [0.7-1.1] | -0.78 | 0.434 |
| FS, median [IQR] | 34 [30-38] | 36 [31-38] | -1.31 | 0.190 |
| EF, median [IQR] | 63 [58-68] | 65 [59-68.25] | -1.22 | 0.221 |

### 3.2 Model construction

The results of COX univariate analysis and LASSO regression analysis on the dependent variables composite endpoint and HF hospitalization showed that the number of predictors could be reduced to 16 and 10, respectively. Taking death as the dependent variable, 19 predictors were identified as independent variables after univariate analysis (Supplementary Tables 1 and 2). To further control for the influence of confounding factors, multivariate COX stepwise regression analysis was performed on the above variables, and a risk prediction model was constructed. The composite endpoint prediction model contained seven variables. A higher number of hospitalizations due to HF (pulmonary rales, hyponatremia, or hypoalbuminemia), previous history of cerebral hemorrhage, aortic or mitral valve calcification, on by echocardiography, and higher NT-proBNP levels could predict the composite outcome (Table 2). The model for predicting HF hospitalization included six variables: older age, previous hospitalizations for HF, hyponatremia at the time of HF, higher NT-proBNP and high-sensitivity troponin T (hs-TnT) levels, higher MVe values measured by cardiac ultrasound could predict HF hospitalization (Table 2). The predictive death model contained seven variables. Longer dialysis age, previous history of atrial fibrillation, presence of aortic or mitral valve calcification (VC) on echocardiography, the occurrence of HF with infection and pleural effusion, low serum sodium levels, and higher hs-TnT could predict death (Table 2 and Supplementary Table 3).

**Table 2.**
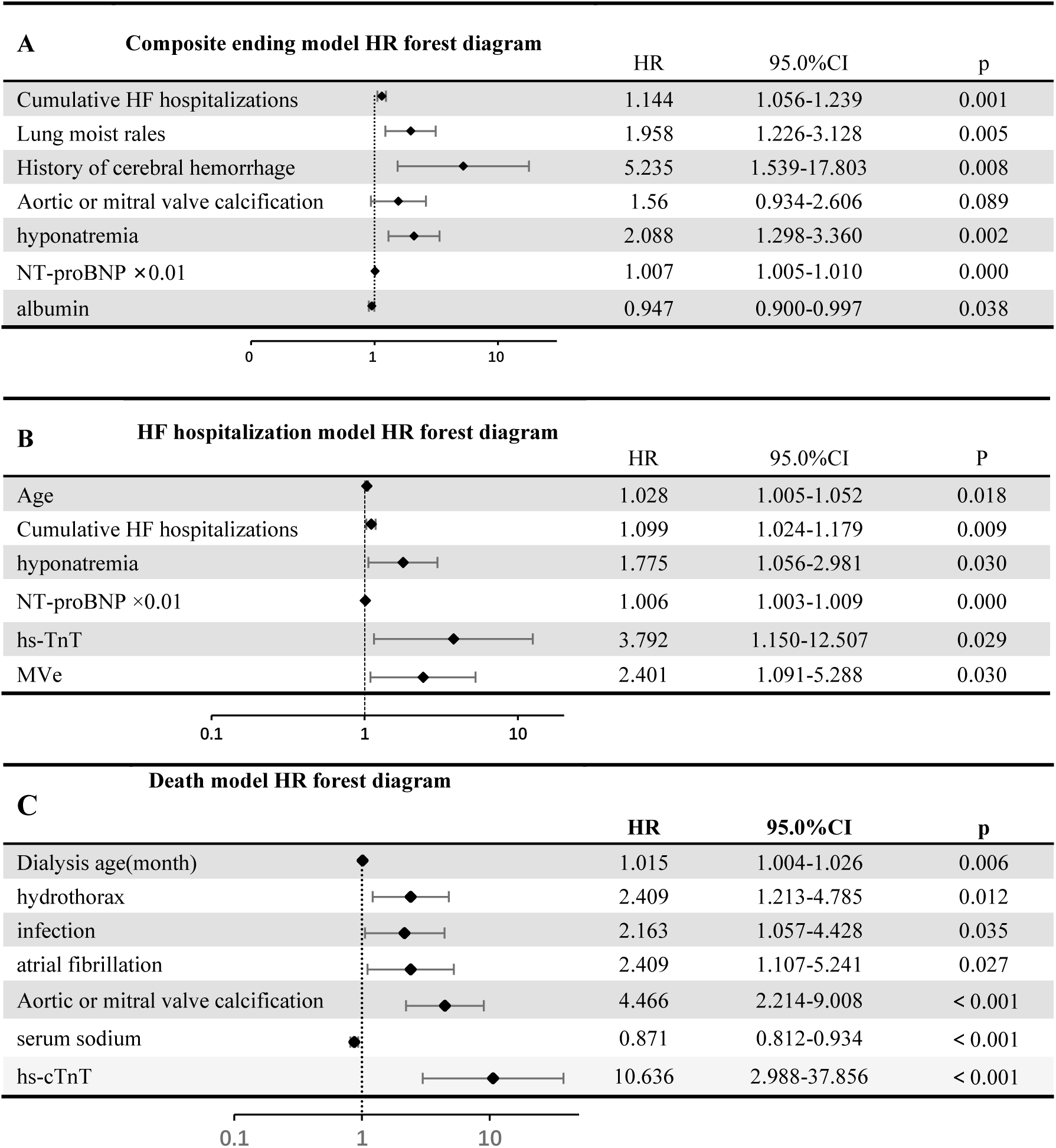
Univariate model built based on the composite outcome

### 3.3 Model verification

According to the composite outcome prediction model, a nomogram was drawn to predict the composite endpoint at 1-2 years. Drawing a line directly to the 1-2–year probability axis after summing the points for each variable to obtain total score, indicates the probability of HF hospitalization or death in 1-2 years. The receiver operating characteristic (ROC) curves of the modeling set and the validation set were plotted, and the area under the curve and C statistics (Table 3) were calculated. The calibration curve, decision curve analysis (DCA), and Kaplan-Meier (K-M) curves were plotted (Figure 2).

**Figure 2:**
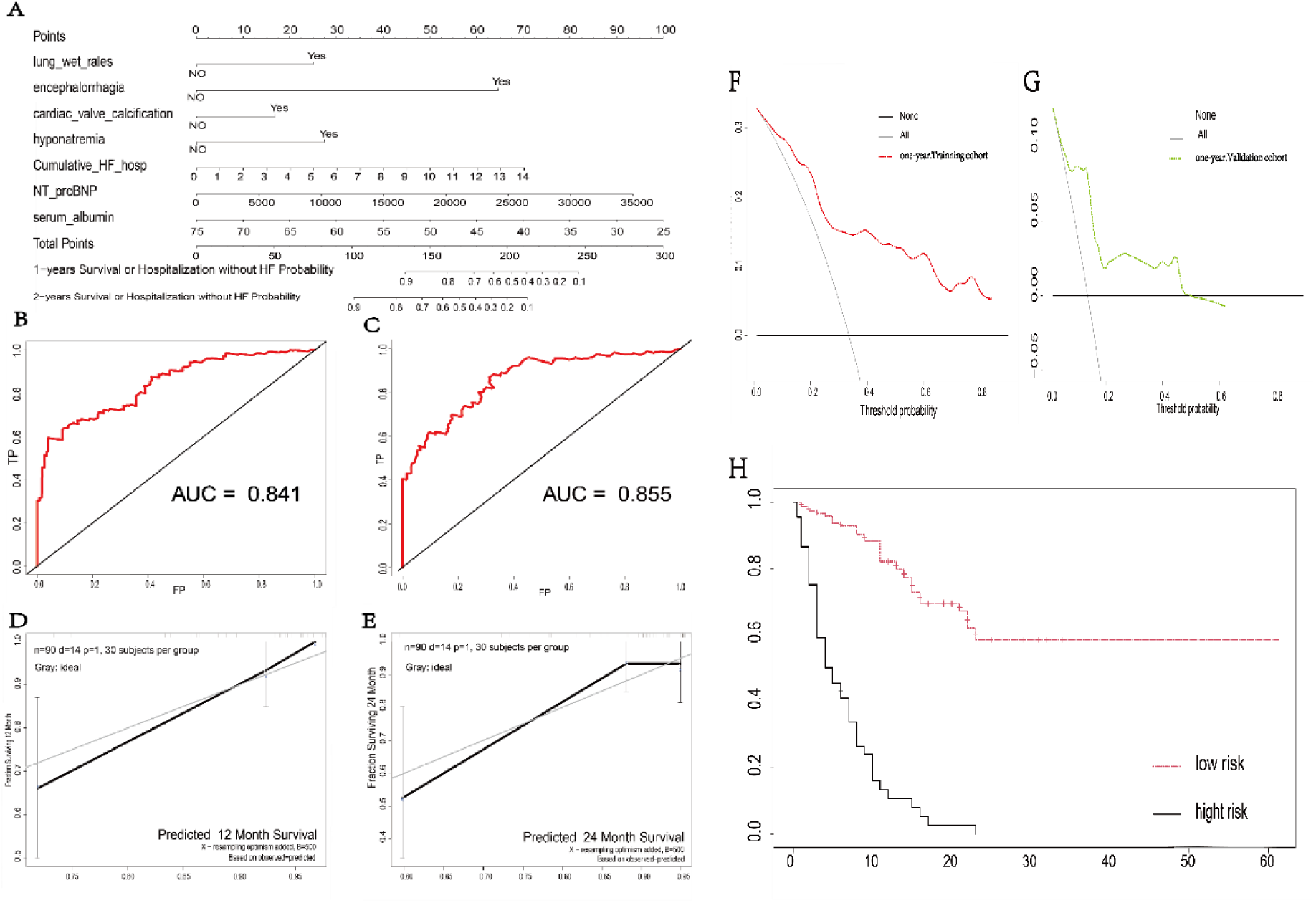
Validation of composite outcome prediction model. (A) Predictive composite outcome model and nomogram. (B-C) The AUC curve of a 1-year and 2-year composite outcome. (D-E) Calibration chart for predicting 1- and 2-year composite outcomes. Clinical decision curve analysis of the nomogram: the Y axis represents fraction surviving, the X axis represents the threshold probability, the red line represents the net benefit of the nomogram model in the training cohort, and the green line represents the net benefit of the nomogram model in the validation cohort. (F) Clinical decision curve analysis of the nomogram for the training cohort and (G) for the validation cohort. The Kaplan–Meier survival curve was drawn according to the risk of death or hospitalization due to HF.

**Table 3.**
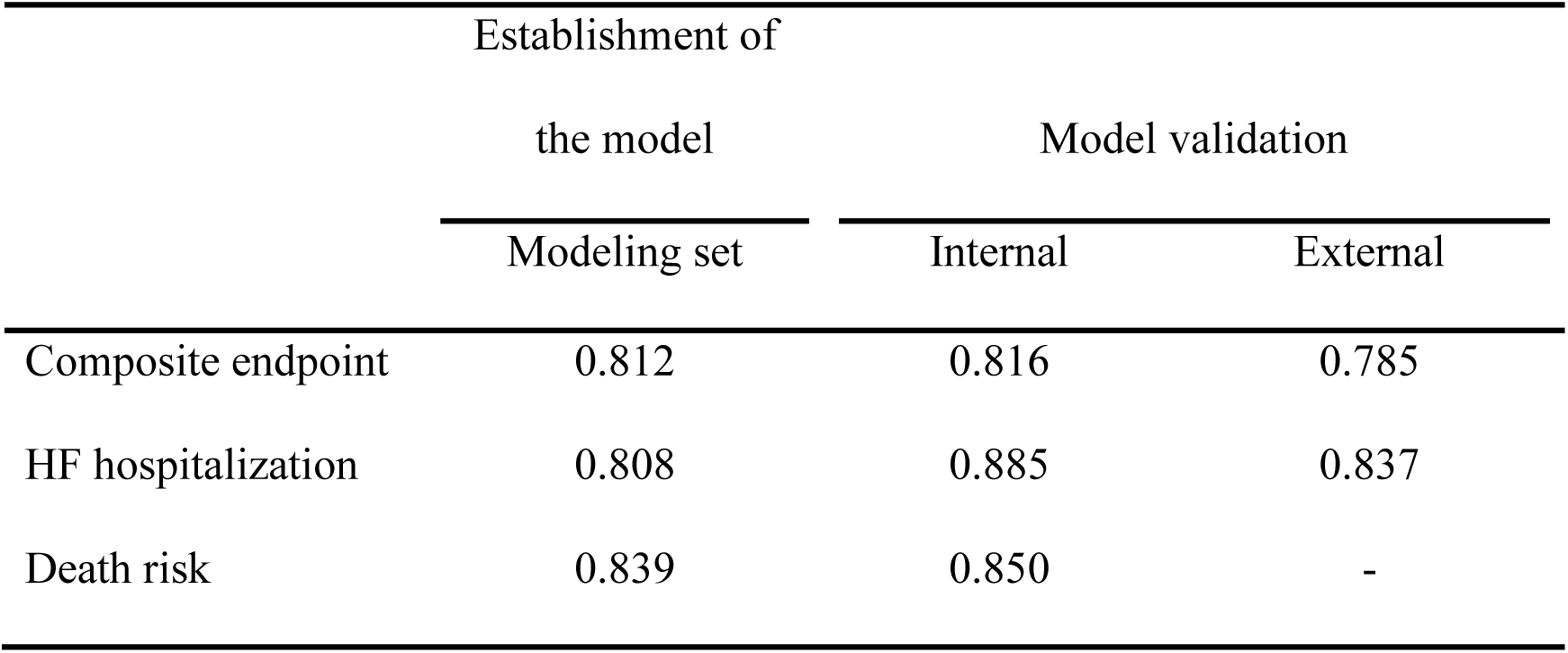
C-statistic values of composite endpoint, HF hospitalization and death model

A nomogram for predicting HF hospitalization was drawn based on the results of the HF hospitalization prediction model. Drawing a line to the 1-2–year probability axis after summing the points of each variable to obtain a total score, indicates the 1-2 year survival probability following HF hospitalization. The receiver operating characteristic (ROC) curves of the modeling set and the validation set were plotted, and the area under the curve and C statistics (Table 3) were calculated. The calibration curve, decision curve analysis (DCA), and Kaplan-Meier (K-M) curves were plotted (Figure 3).

**Figure 3:**
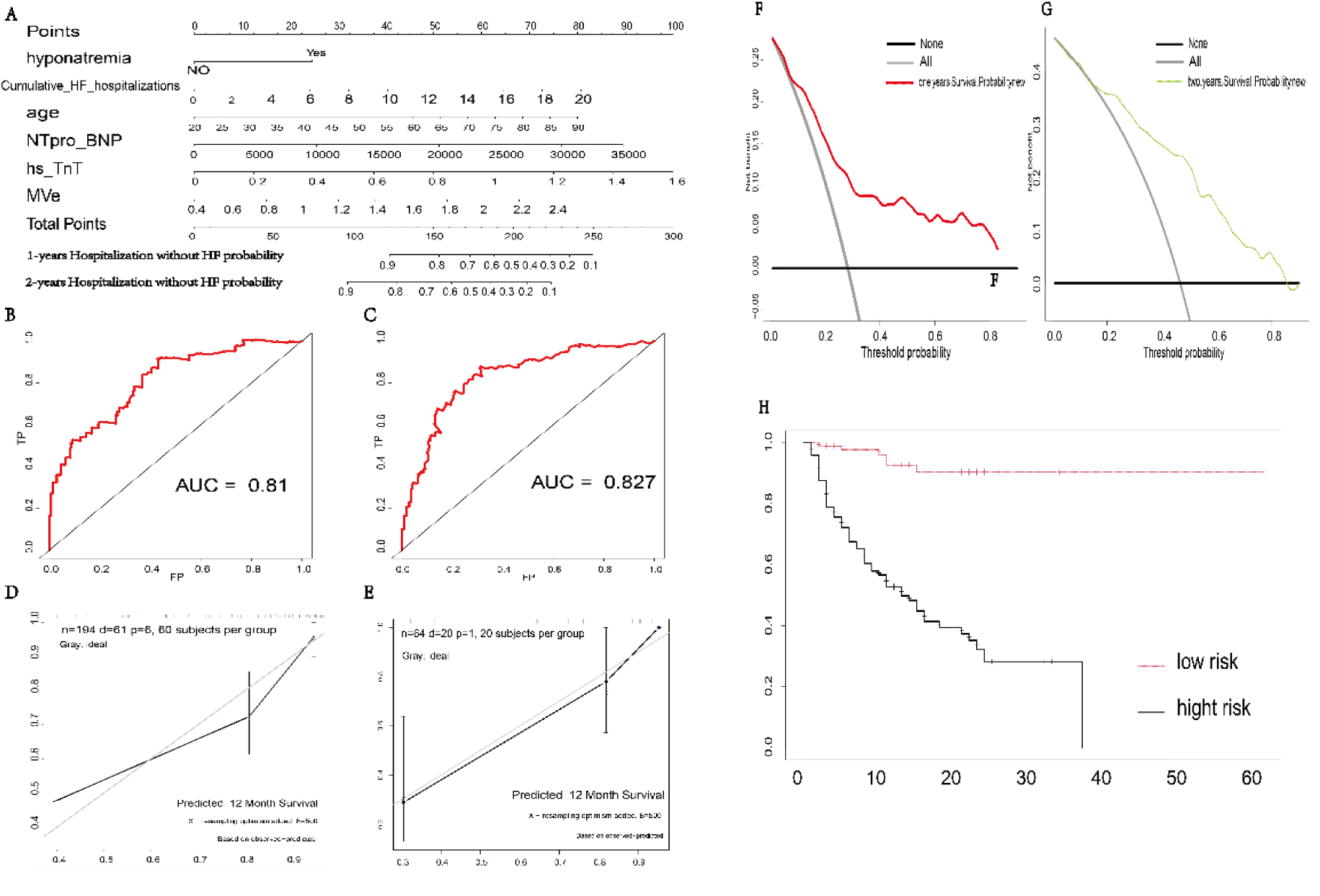
HF hospitalization prediction model validation. (A) Nomogram for predicting heart failure (HF) hospitalization. (B-C) AUC curve predicting a 1-year HF hospitalization. (D-E) The 1-year heart failure outcome calibration chart. Clinical decision curve analysis of the nomogram The Y axis represents the fraction surviving; the X axis represents the threshold probability; the red line represents the net benefit of the nomogram model in the training cohort; the green line represents the net benefit of the nomogram model in the validation cohort. (F) Clinical decision curve analysis of the nomogram in the training cohort and (G) in the validation cohort. (H) Kaplan–Meier survival curve according to the risk of HF hospitalization.

Based on the death outcome prediction model, a nomogram was drawn to predict 1–2 year death outcomes. After the summing the points of the individual variables this correspond to a total score on the total points axis, which is issued to indicated the probability of 1-2-year death by drawing a line to the 1-2 year survival probability axis. The receiver operating characteristic (ROC) curves of the modeling set and the validation set were plotted, and the area under the curve and C statistics (Table 3) were calculated. The calibration curve, decision curve analysis (DCA), and Kaplan-Meier (K-M) curves were plotted (Figure 4).

**Figure 4:**
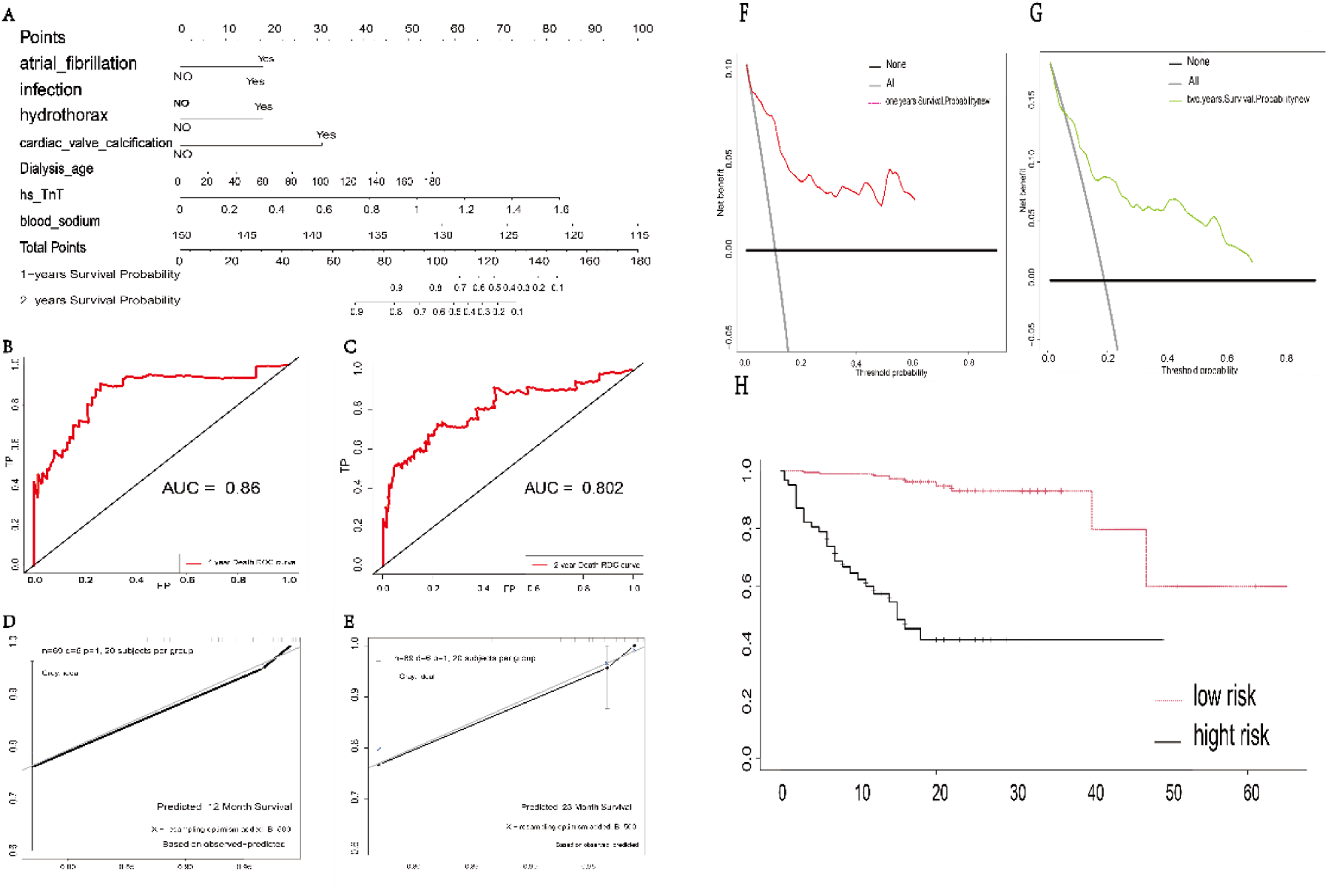
Verification of death prediction model. (A) Nomogram for predicting death. (B-C) AUC curve for the 1-year composite outcome. (D-E) Calibration plot for predicting 1- and 2-year heart failure outcomes. Clinical decision curve analysis of the nomogram. The Y axis represents the fraction surviving; the X axis represents the threshold probability; the red line represents the net benefit of the nomogram model in the training cohort; the green line represents the net benefit of the nomogram model in the validation cohort. (F) Clinical decision curve analysis of the nomogram for the training cohort and (G) for the validation cohort. (H) Kaplan-Meier survival curve according to the risk of HF hospitalization.

COX regression analysis and 10-fold cross validation were performed on the training cohort and the internal and external validation sets. The results showed that the AUC values (Figures 2–4) and c-statistics of the three prediction models were remained above 0.8 (Table 3), indicating the model’s discrimination ability was good. For both internal and external validation cohort, the calibration plot showed a good consistency between the predicted HF hospitalization, mortality risk, and actual risk of the nomogram. The DCA curve indicated that the prediction model © had good clinical usability. The respective Kaplan-Meier survival curves showed that the three models could effectively distinguish high-risk and low-risk groups (Figures 2–4).

We use variables in the compact model for risk scoring. The decision tree algorithm performs the best classification according to the critical points shown in the figure. For each ‘bad’ variable, the patient increases the score by 1 point. The score range for mortality, HF hospitalization rate, and the combined endpoint is 0-3. Finally, the corresponding K-M curve is drawn according to the score. (Figure 5).

**Figure 5.**
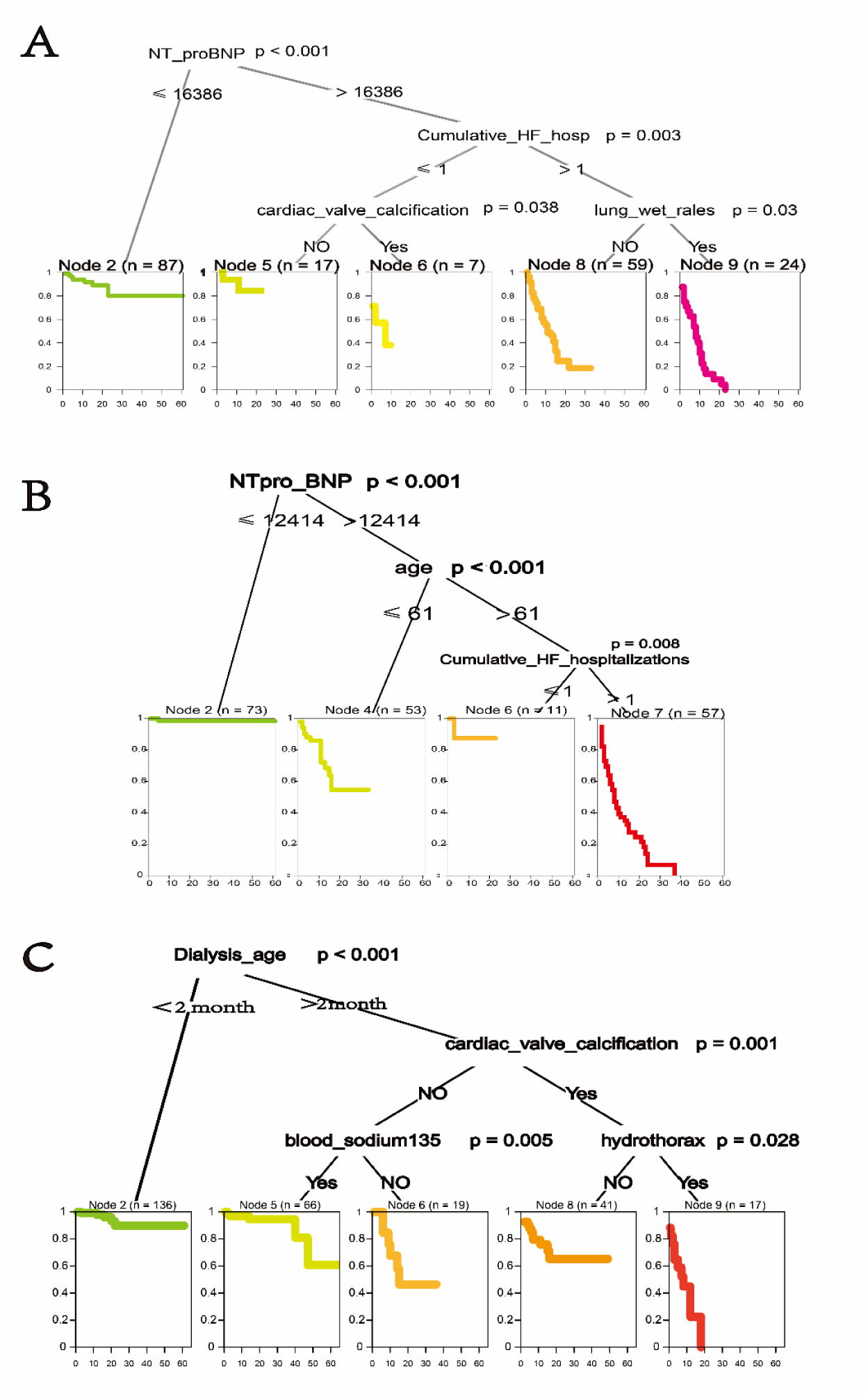
(A) composite outcome prediction model; (B) heart failure hospitalization prediction model; (C) death prediction model

## 4. Discussion

HF is a common cardiovascular disease in MHD patients, which significantly increases the hospitalization rate, mortality, and poses significant economic burden on MHD treatment. However, it is still difficult to predict the mortality and hospitalization rate of MHD patients with HF^[4–6]^. Most of the published prediction models have not been externally validated and are not suitable for MHD patients. We screened different variables using 112 predictors by LASSO regression and multivariate COX regression analysis and constructed a prediction model with HF hospitalization, death, and composite outcome as outcome events. To our knowledge, this study established the first model suitable for the prediction of HF risk in patients receiving MHD. The c-statistics of the internal and external validation sets were >0.8, indicating that the model’s prediction performance was good. The modeling dataset and the verification dataset were completely independent cohorts obtained from different centers, which verifies the applicability and stability of the prediction model. Secondly, because patients receiving MHD will regularly attend a hemodialysis center for treatment, we obtained follow-up information relatively accurately, thus ensuring the authenticity and reliability of the data obtained. In addition, the predictive model provided in this study can provide effective prognostic information for patients receiving MHD with only a small number of easily available clinical variables, indicating that the model is suitable for the real-life clinical setting and is convenient to use.

With regard to the choice of diagnostic cutoff value for NT**-**proBNP levels, there are currently different viewpoints. First, in patients undergoing MHD, the glomerular filtration rate (GFR) is reduced, NT-proBNP clearance rate decreases, and volume overload occurs. This increased volume state causes BNP to be secreted in conjunction with increased myocardial wall relaxation. Second, a cutoff of NT-proBNP <125 pg/mL (negative exclusion) has been proposed by the latest guidelines in Europe and the United States for common HF or NT-proBNP levels >2000 pg/mL have been established by similar studies, but are not applicable to HF in patients receiving MHD. In contrast, the study by Jafri et al. suggested that the sensitivity and specificity of NT-proBNP values ≥112,15.2 pg/mL for the diagnosis of HF in patients with CKD5 were 94.7% and 100%, respectively^[19]^. In addition, in our study, we also diagnosed HF using the above three cut-off values and constructed a diagnostic model and found that using the NT-proBNP <125 pg/mL or >2000 pg/mL cutoff values led to overdiagnosis of HF. Furthermore, the model constructed using the criteria of NT-proBNP >11,215.2 pg/mL had better predictive power (Figure S3). In conclusion, NT-proBNP >11,215.2 pg/mL was selected as the cutoff value for the diagnosis of HF in patients receiving MHD patients for this study.

Interestingly, the predictors of the three prediction models partially overlapped. In our prediction model, hyponatremia simultaneously predicted the risk of HF hospitalization and death in MHD patients. Studies in the general population have shown that hyponatremia is a good predictor of adverse outcomes in acute or chronic HF morbidity and mortality^[14,20,21]^. The study by Marroquin et al. showed that hyponatremia was associated with CVD before the transition to end-stage kidney disease (ESKD) and a high hospitalization rate after ESKD transition^[22]^. The results of this study further indicated that hyponatremia is still an important predictor of HF hospitalization and composite outcomes in MHD patients. Although the predictive factor included in the mortality prediction model is serum sodium, it also indicates that lower serum sodium is associated with a higher risk of death. However, it is still not clear whether hyponatremia directly leads to higher hospitalization rates and mortality or is a sign of potential complications (such as severe infection or lung disease). Recent studies by Marroquin et al. found that there is still a close relationship between hyponatremia and higher mortality and hospitalization rates after adjusting for comorbidities^[22]^. We also obtained similar results after adjusting for comorbidities in our models (Supplementary Table 4).

Our study found that NT-proBNP appeared in both the HF hospitalization and composite outcome prediction models, while hs-cTnT appeared in both HF hospitalization and death risk prediction models. A recent meta-analysis found that NT-proBNP serum levels in patients with ESKD were positively correlated with cardiovascular events and mortality risks^[23]^. In addition, we also used the best decision tree method to determine that NT-proBNP >16,386 pg/mL and >12,414 pg/mL were high-risk groups for compound outcomes and HF hospitalization in MHD patients, respectively (Figure 5). Clinically, hs-cTnT is most commonly used to determine the presence of myocardial infarction or HF-related injury in patients with acute HF (AHF). However, in the absence of myocardial ischemia, the elevation of hs-cTnT indicates the presence of strong myocardial cell injury, progressive ventricular remodeling, and an increased risk of death^[24]^. Chesnaye et al. found that high levels of hs-cTnT predicted a higher risk of death and a poorer prognosis in patients with stage 4-5 CKD^[25]^. This study shows that hs-cTnT is an independent predictor of death in MHD patients, and the combination of NT-proBNP and hs-cTnT could effectively predict the risk of HF hospitalization in these patients. Interestingly, our study also found that cardiac VC was a predictive factor in both the death and compound outcome models. The study found that the prevalence of aortic and mitral valve VC in MHD patients was eight times higher than that in the general population^[26–28]^. In patients receiving MHD, hyperphosphatemia, hyperparathyroidism, increased fibroblast growth factor-23, and decreased co-receptor klotho levels are significantly associated with VC^[27]^. Cardiac VC can lead to impaired valve opening and closing function and then rapidly progresses to valve stenosis. In particular, aortic valve stenosis increases blood flow resistance, causing left ventricular hypertrophy and HF leading to premature death^[26,27]^. In summary, compared with the prediction model for HF in the general population, VC has a unique and good predictive value for predicting death in patients receiving MHD.

It is noteworthy that there were also significant differences in the predictors for different models. Low albumin, pulmonary rales, and a history of cerebral hemorrhage only appeared in the composite outcome prediction model, while age and MVe only appeared in the HF hospitalization model. Although E/A and E/e ratios have been repeatedly used to predict cardiovascular death and HF hospitalization in previous studies, they were not included in our model analysis due to the large differences in ultrasound reports collected from various centers^[29]^. Surprisingly, this study also effectively predicted the risk of HF hospitalization in MHD patients through the simple MVe value in echocardiography, which seemed to increase the general applicability of the model. In addition, we also found that atrial fibrillation (AF), infection, dialysis age, and pleural effusion only appeared in the death prediction model. A number of studies have shown that AF is closely related to HF, which significantly increases the incidence and mortality due to CVD in MHD patients. Early screening and intervention in patients with AF can effectively improve the poor prognosis of high-risk groups^[30–34]^. Recently, Evangelos et al. together with the numerical citation.proposed that the factors hyponatremia, AF, and HF constitute a difficult triangle, and any side of it will affect the other side, increasing morbidity and mortality^[35]^. The death prediction model established in this study includes these three important factors, which shows that our model is suitable for the real-world clinical setting. Pleural effusion has rarely been mentioned in previous predictive models or mechanism studies. In this study, the results of logistic multivariate analysis showed that the formation of pleural effusion may be closely related to respiratory failure, pulmonary edema, HF, pulmonary infection, and AF (Supplementary Table 5), and that pleural effusion may reflect the existence of severe complications in MHD patients and may predict the occurrence of death. It is expected that future research will demonstrate its correlation with death outcomes and its mechanism.

Our research has some limitations that should be considered. First, this study was a multi-center retrospective cohort study. The clinical data derived from four hospitals in different regions. The health care system and patient treatment vary greatly, which may affect management, outcomes, and prediction. Second, in order to ensure the integrity of the data, we only collected the patient’s previous hospitalization data and did not collect data with mild symptoms or remission after outpatient treatment, which may have led to a selection bias in the data, but nonetheless allowed our model to distinguish the HF hospitalization risk. Compared with prospective randomized controlled trials, we failed to include variables that are currently a focus of research attention into the analysis (such as biomarkers, genomics, and proteomics information) and did not effectively control for confounding factors. Finally, the outcome was determined by the attending physician alone and not by a ruling committee. Although, a systematic meta-analysis did not find any effect due to differences in event adjudication on the conclusions of cardiovascular outcome trials^[36]^.

## 5. Conclusion

This study developed and validated models for predicting mortality, HF hospitalization, and composite outcomes in patients receiving MHD. The models are suitable for application in MHD patients and for use at different levels of medical institutions. The prediction performances of the models were good, stable, and easy to use. In addition, the prediction models propose a simplified risk score for clinical practice to effectively stratify patients and provide effective prognostic information.

## Institutional Review Board Statement

The study was conducted in accordance with the Declaration of Helsinki, and approved by the Medical Ethics Committee of Nanchong North Sichuan Medical College Second Clinical College (Nanchong Central Hospital) (No: 2022-055 and date of approval 07.18. 2022) for studies involving humans.

## Informed Consent Statement

Informed consent was obtained from all subjects involved in the study.

## Author contributions

All the authors participated in the research and design. Wenwu Tang, Ying Zhang, and Xinzhu Yuan contributed to the statistical analysis. Wenwu Tang, Zhixin Wang, and Xiaoxia Chen provided these data. Wenwu Tang and Zhirui Qi participated in the data interpretation. Ju Zhang and Jie Li drafted the manuscript. Xishengxie and Xiaohua Yang provide supervision or guidance. Each author contributed important knowledge content in the process of drafting or revising the manuscript.

## Data Availability

The data that support the findings of this study are available from the corresponding author upon reasonable request.

## Abbreviations

AST: aspartate aminotransferase
AAO: ascending aorta
ALT: alanine aminotransferase
AO: aorta or aorta root
AV: aortic valve
BMI: body mass index
CKD: chronic kidney disease
COPD: chronic obstructive pulmonary disease
CVD: cardiovascular disease
DCA: decision curve analysis
FS: fractional shortening
HF: heart failure
hs-cTnT: high sensitivity troponin T
IQR: interquartile ranges
IVS: interventricular septum
KDIGO: Kidney Disease Improving Global Outcomes
LA: left atrium
LASSO: Least Absolute Shrinkage and Selection Operator
LV: left ventricle
LVEF: left ventricular ejection fraction
LVPW: left ventricular posterior wall
MHD: maintenance hemodialysis
MPA: main pulmonary artery
MVe: anterior mitral flow velocity
NT-proBNP: N-terminal B-type natriuretic peptide
NYHA: New York Heart Association
RA: right atrium
RLS: 
ROC: construct receiver operating characteristic
RV: right ventricle
TRIPOD: Transparent Reporting of a Multivariate Prediction Model for Individual Prognosis or Diagnosis

## List of supplementary materials

Table S1: Single factor COX analysis

Figure S1: HR values of single-factor COX regression analysis

Figure S2: LASSO Regression analysis

Table S2: Variable screening process

Table S3: Multivariate COX analysisMulti-factor Cox model predictor distribution comparison

Figure S3: Diagnosis of heart failure and construction of logistic models according to different criteria

Table S4: Association of hyponatremia with heart failure hospitalization and death after adjustment for comorbidities

Table S5: A logistics model with pleural effusion as an outcome event and multiple comorbidities as predictors was constructed

---

**Table S1.**
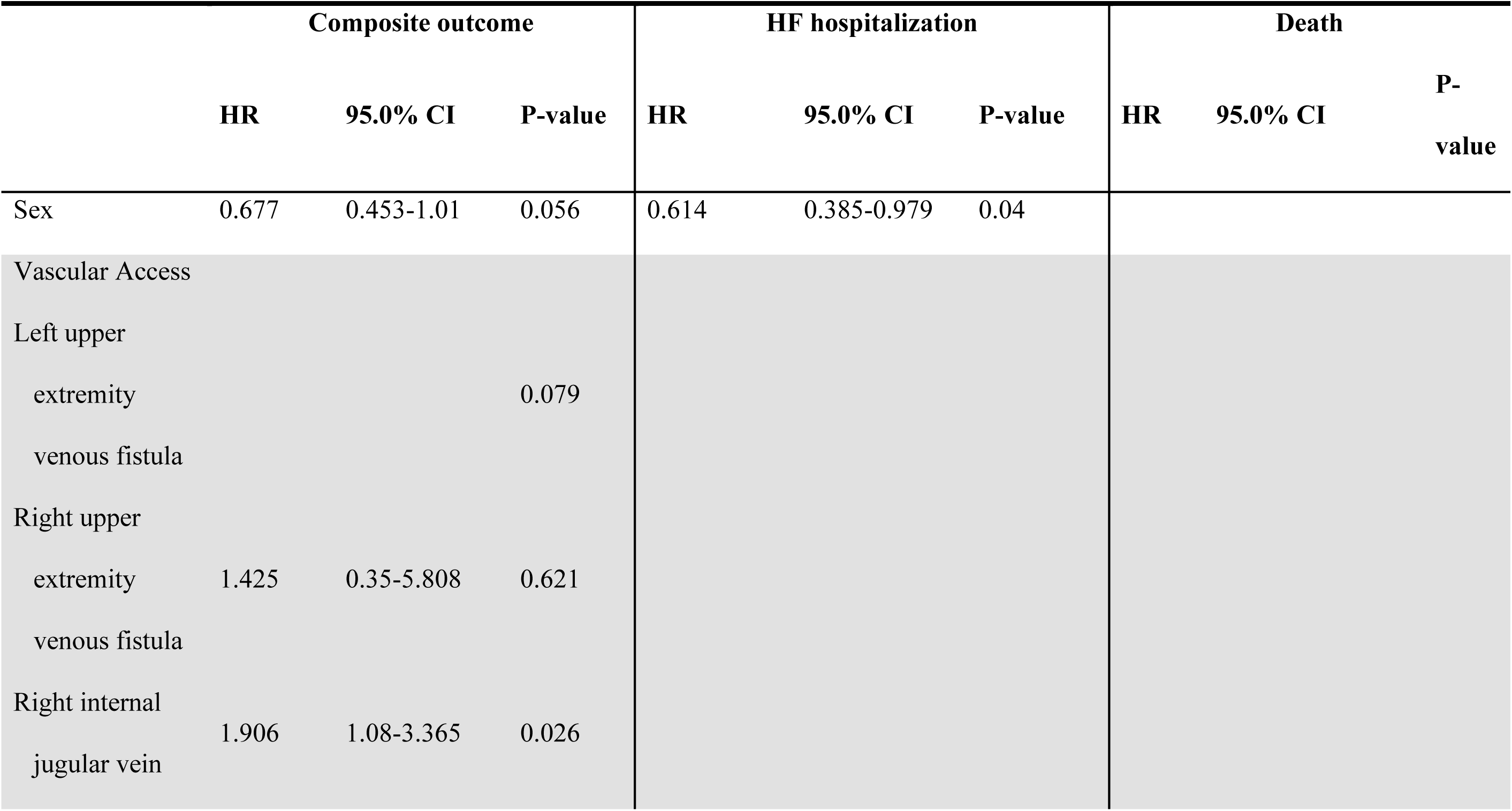
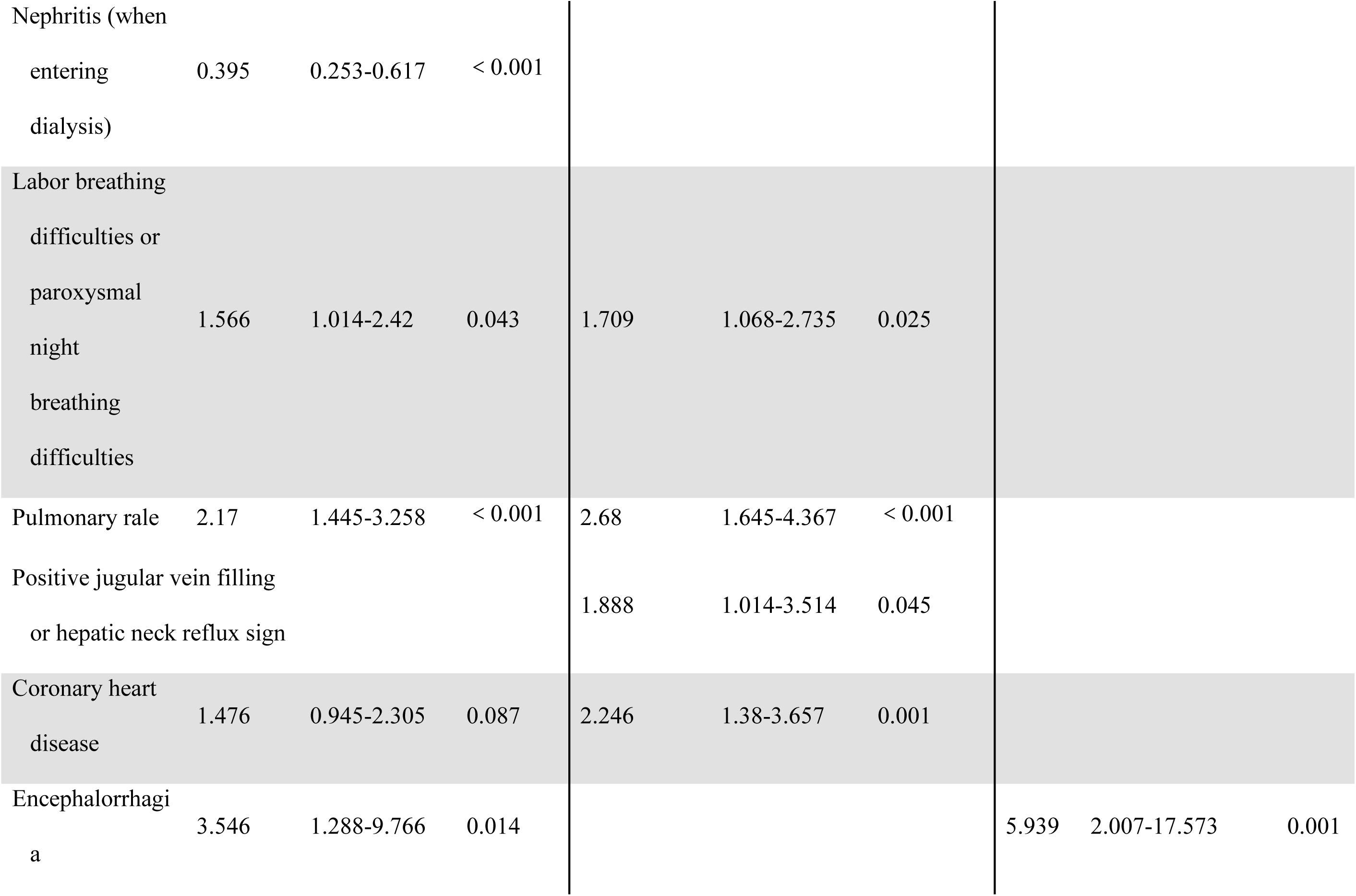
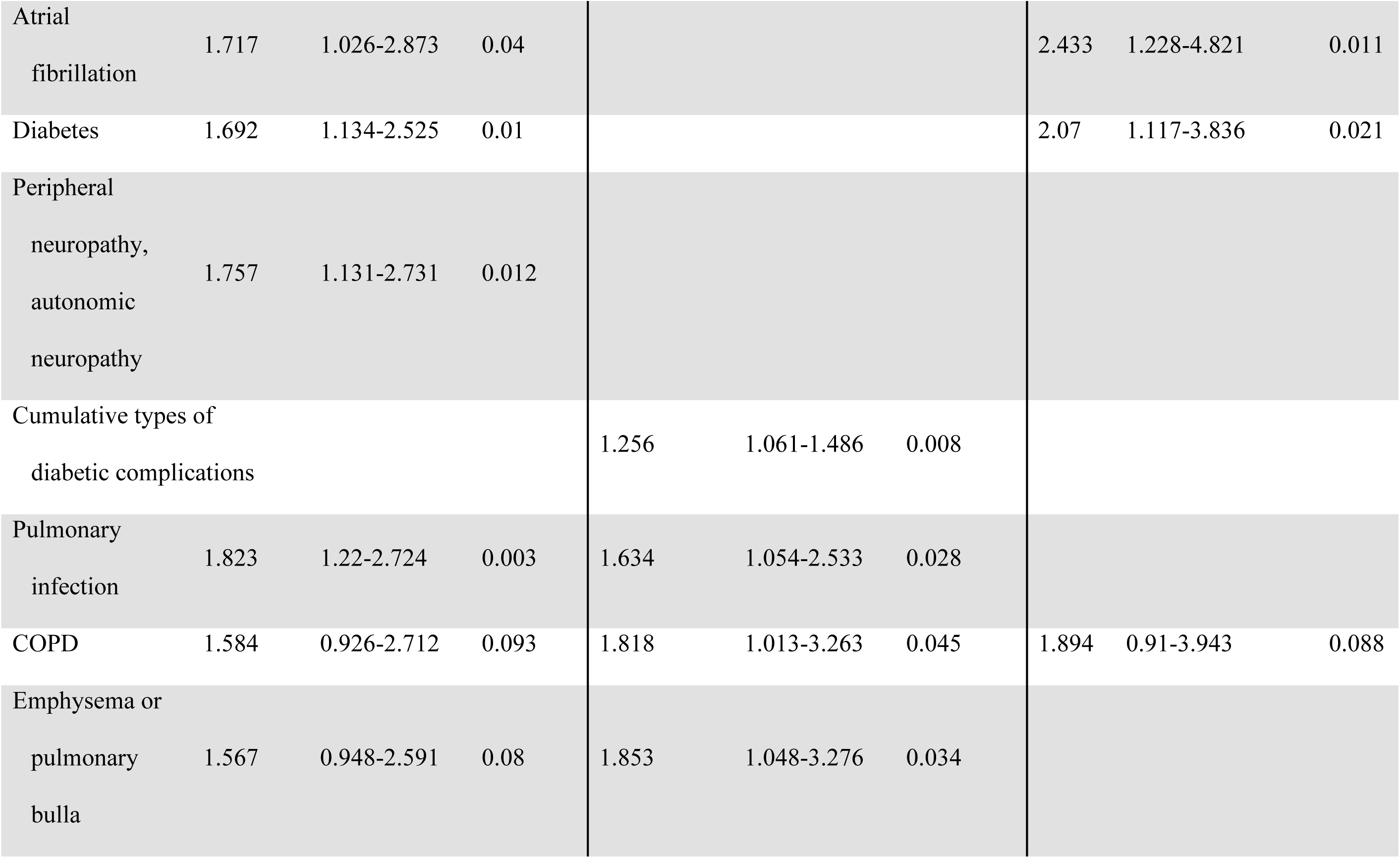
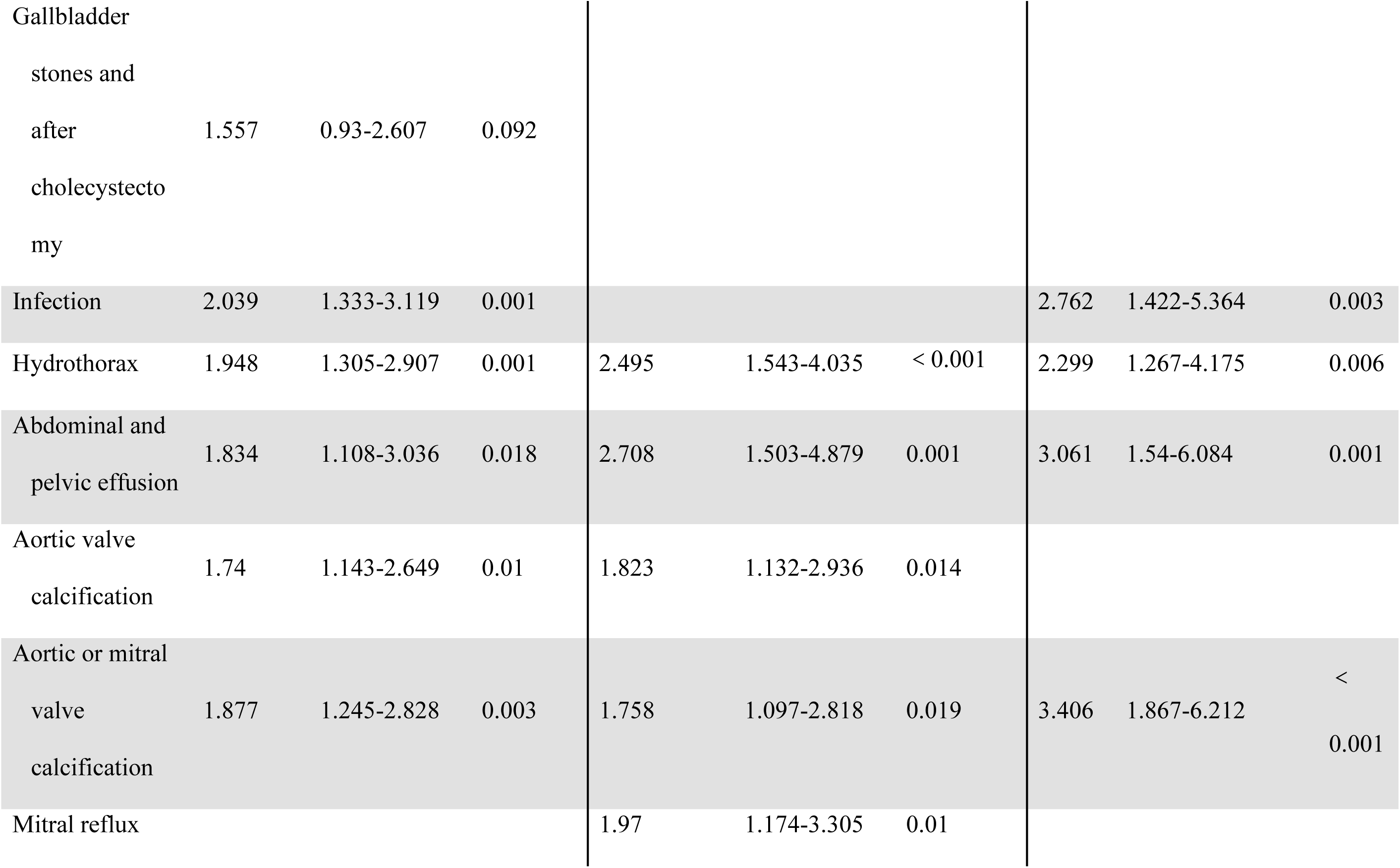
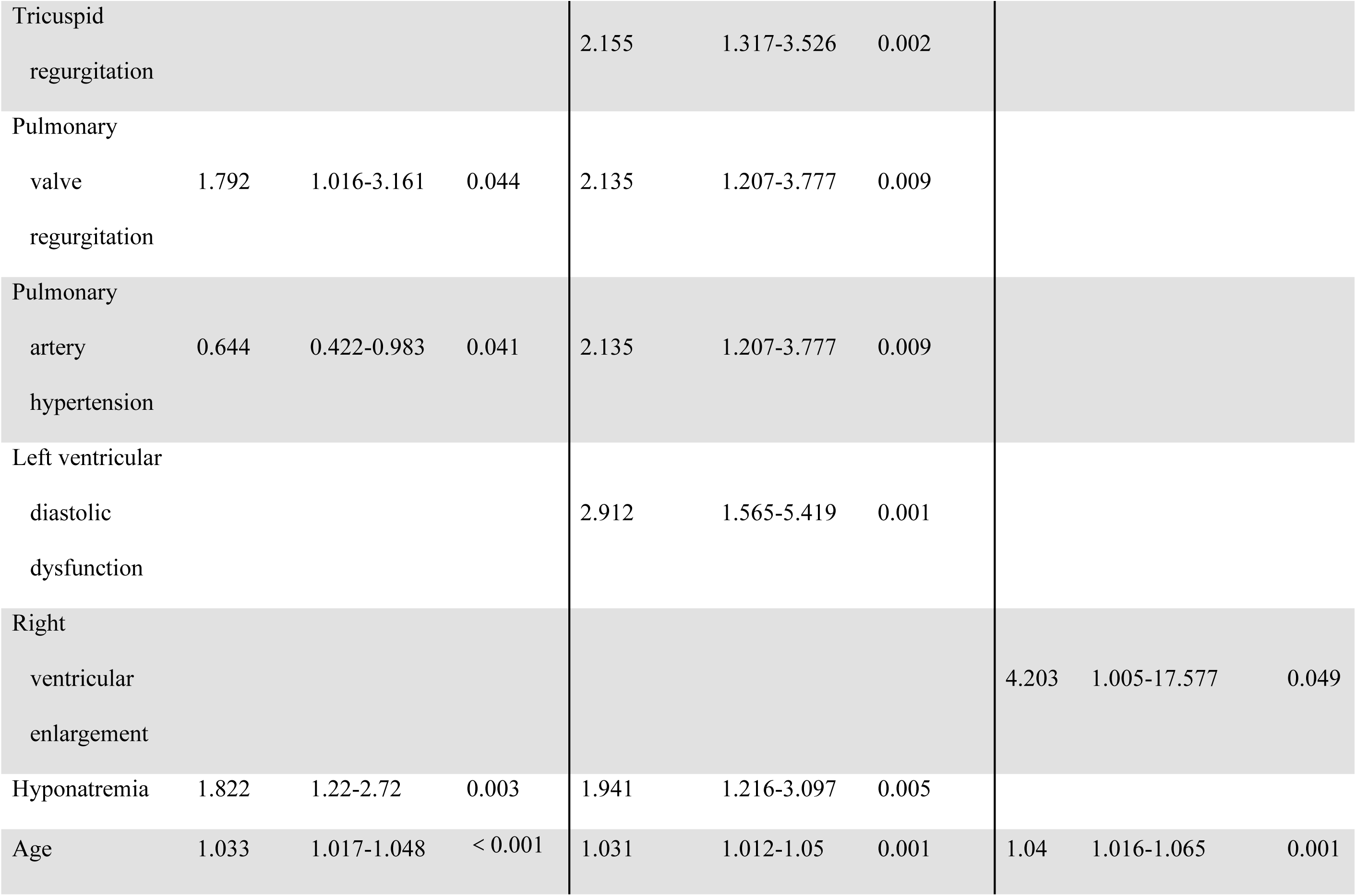
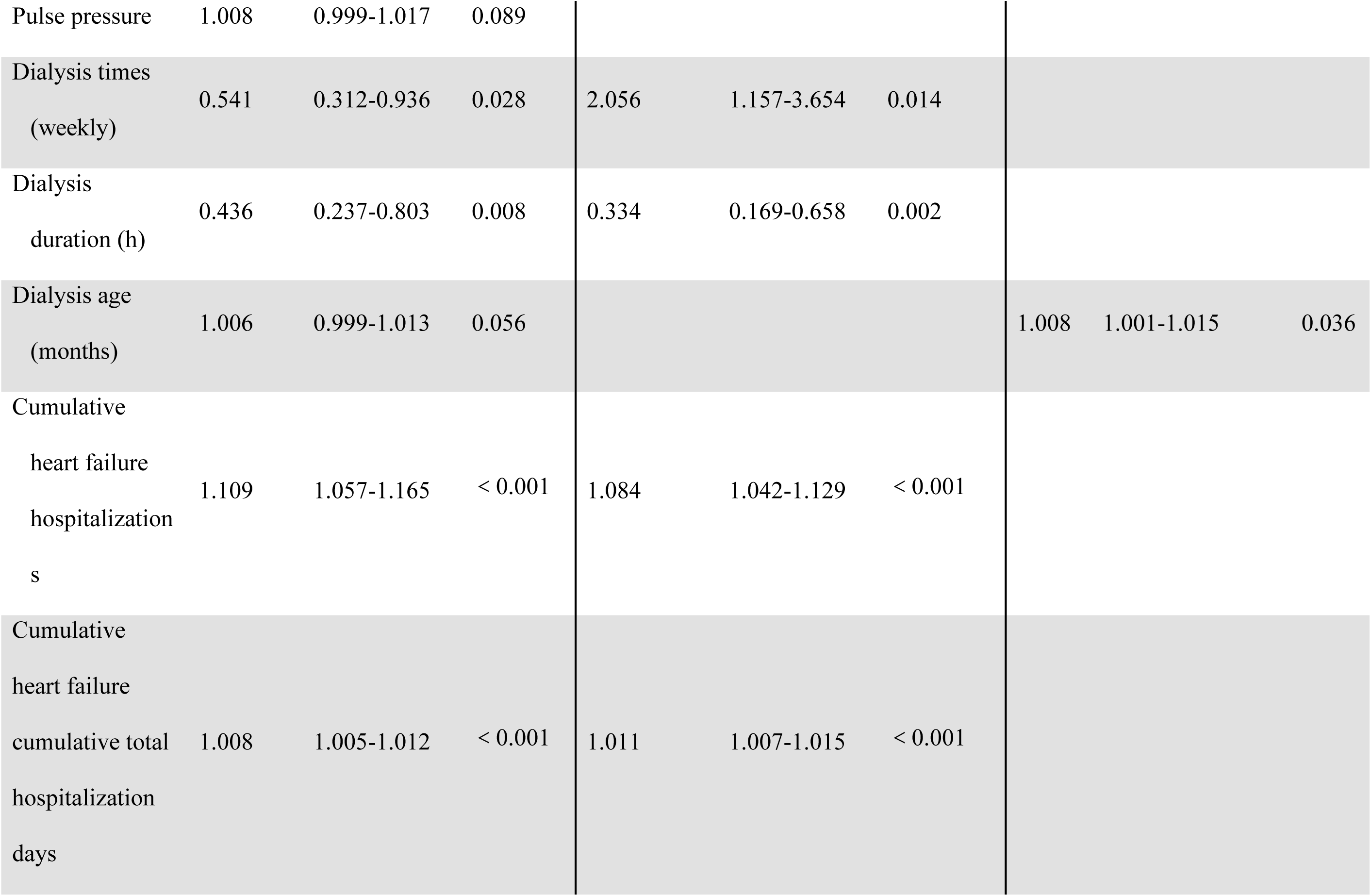
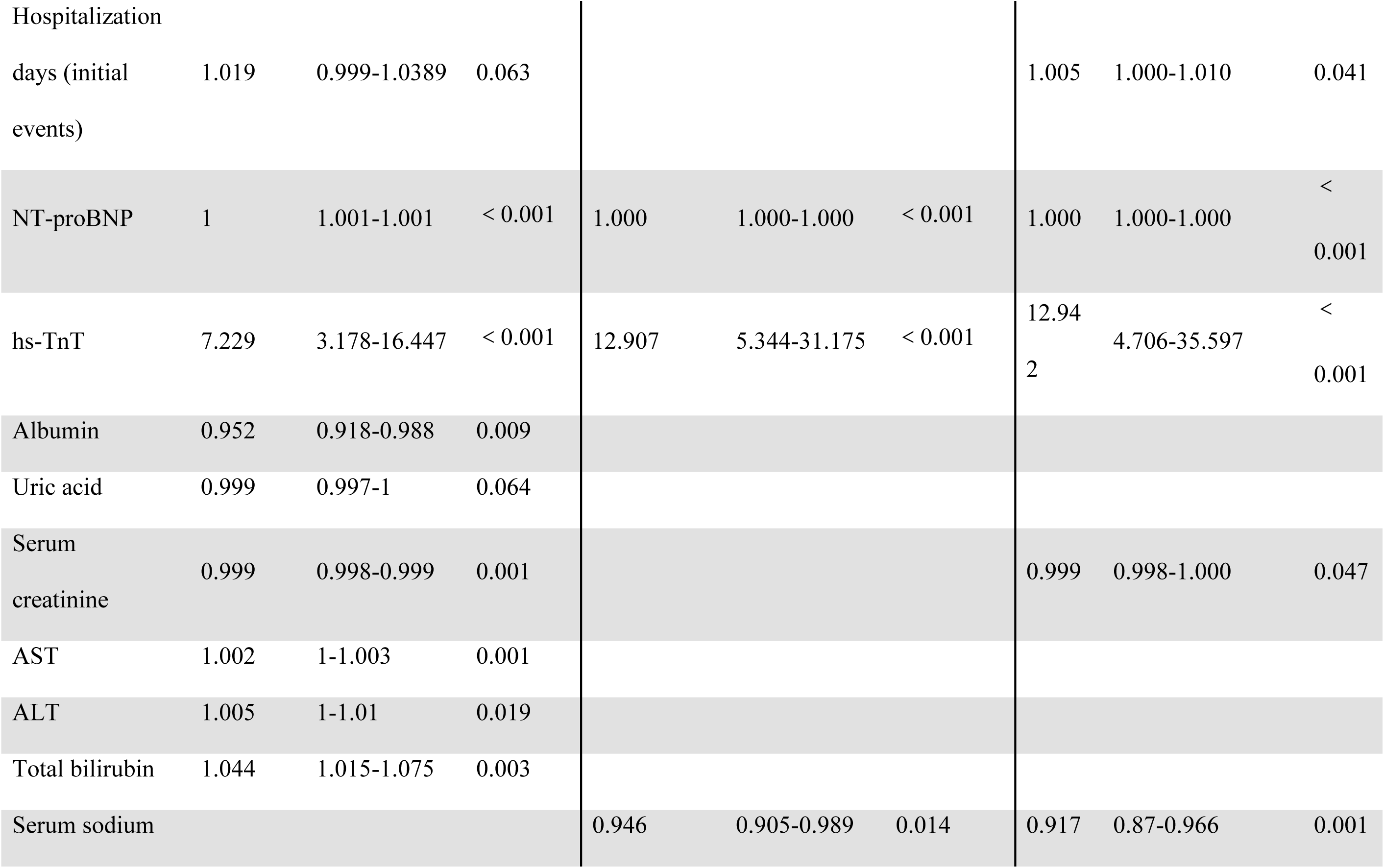
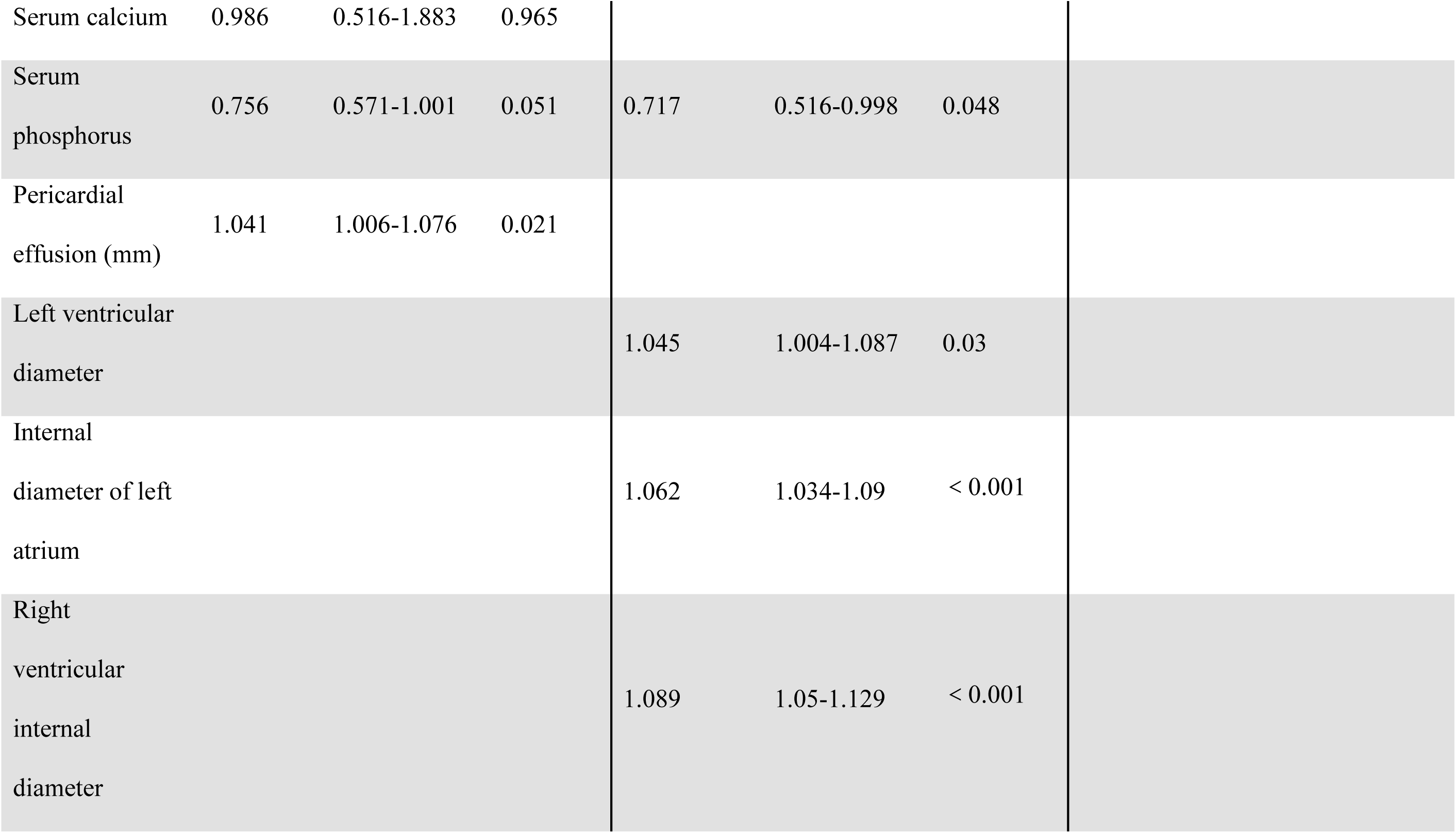
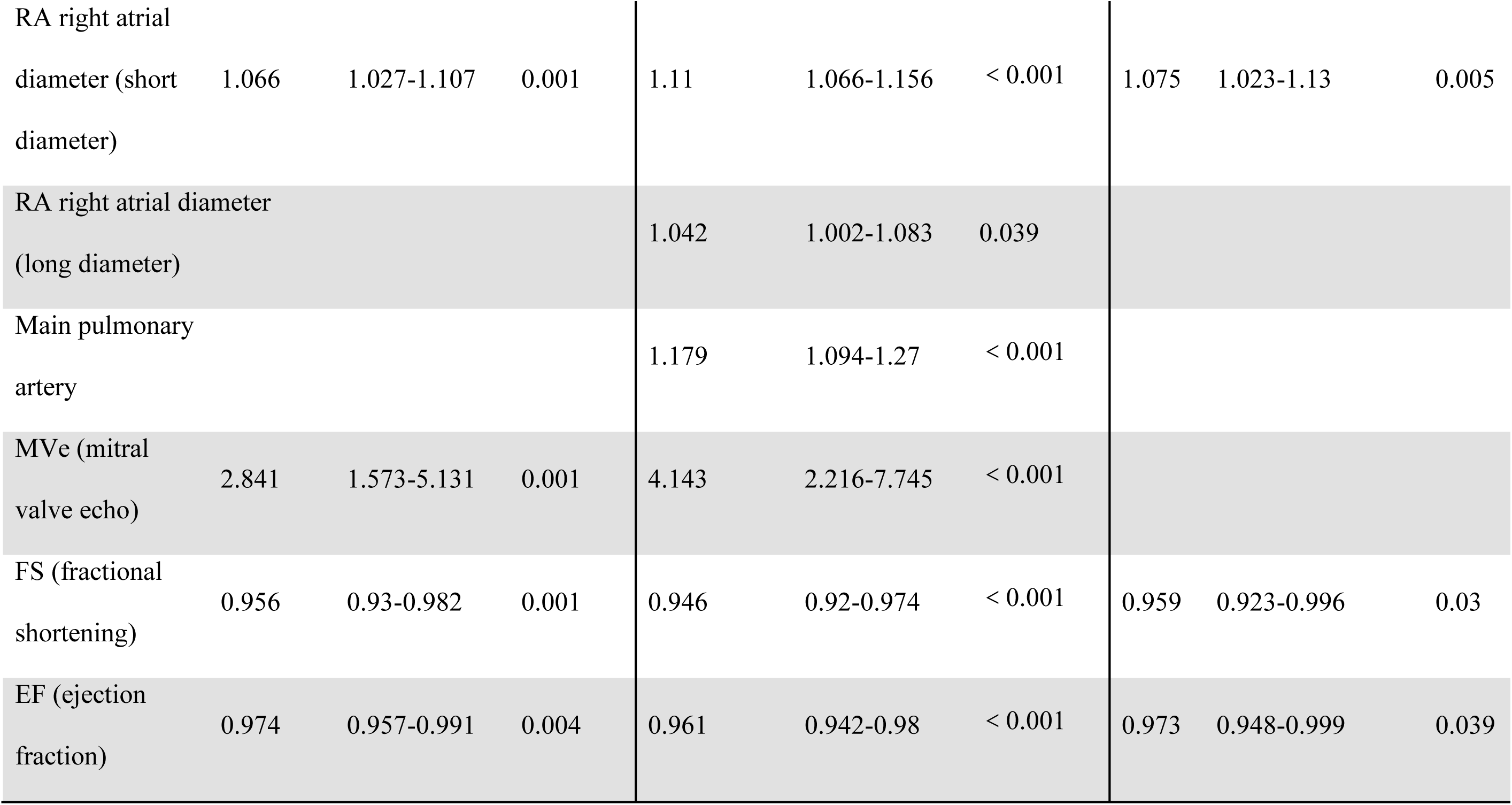
Single factor COX analysis.

**Figure S1:**
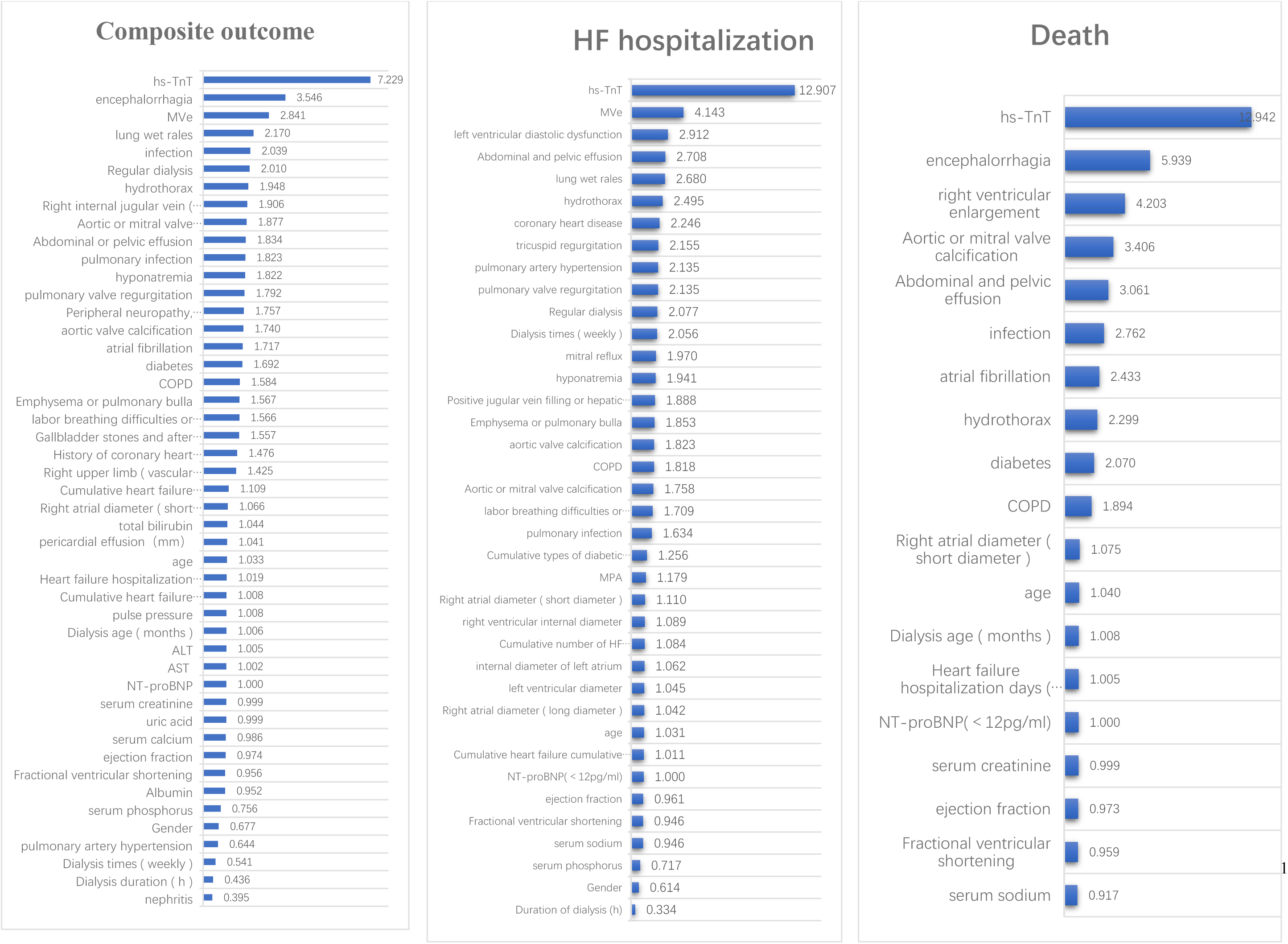
HR values of single-factor COX regression analysis.

**Figure S2:**
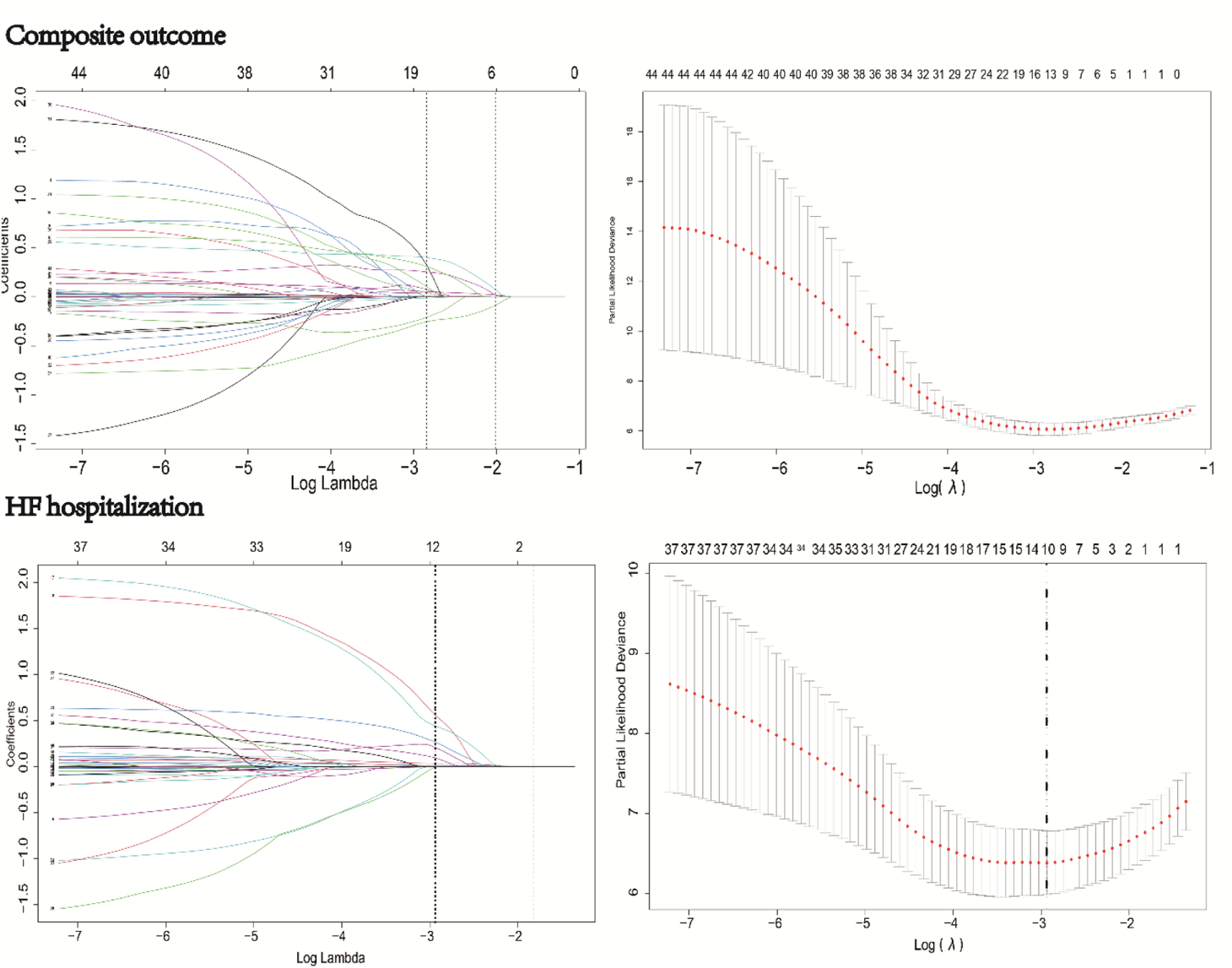
LASSO Regression analysis. **Death outcome:** After single-factor COX analysis, the predictors of death outcome entered the multiple regression equation. A multivariate COX stepwise regression (forward method) was directly performed, and the model was constructed.

**Table S2:**
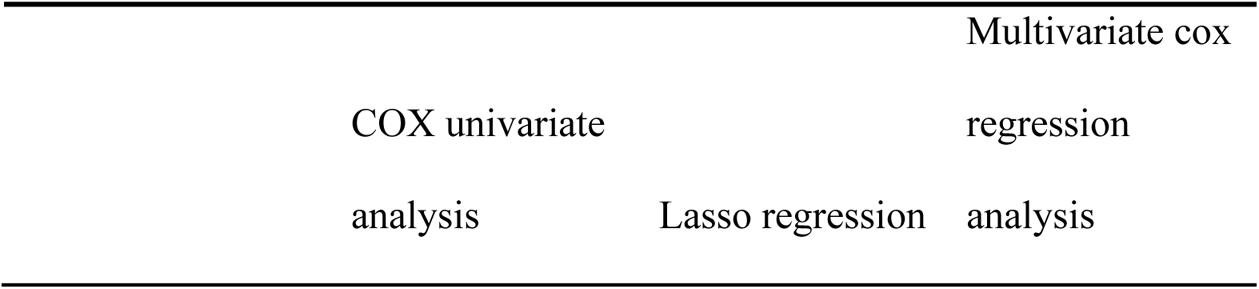
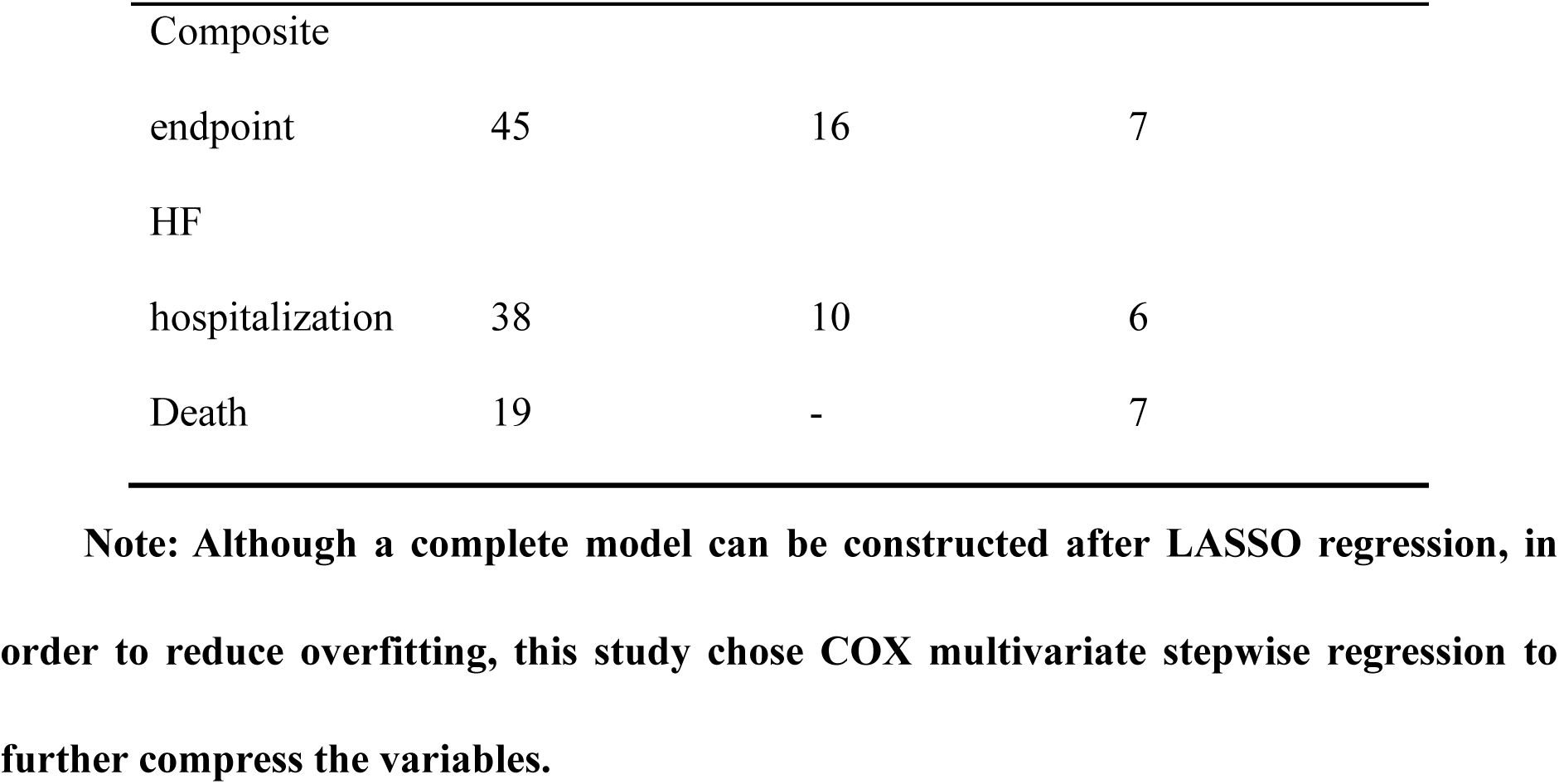
Variable screening process.

**Table S3:**
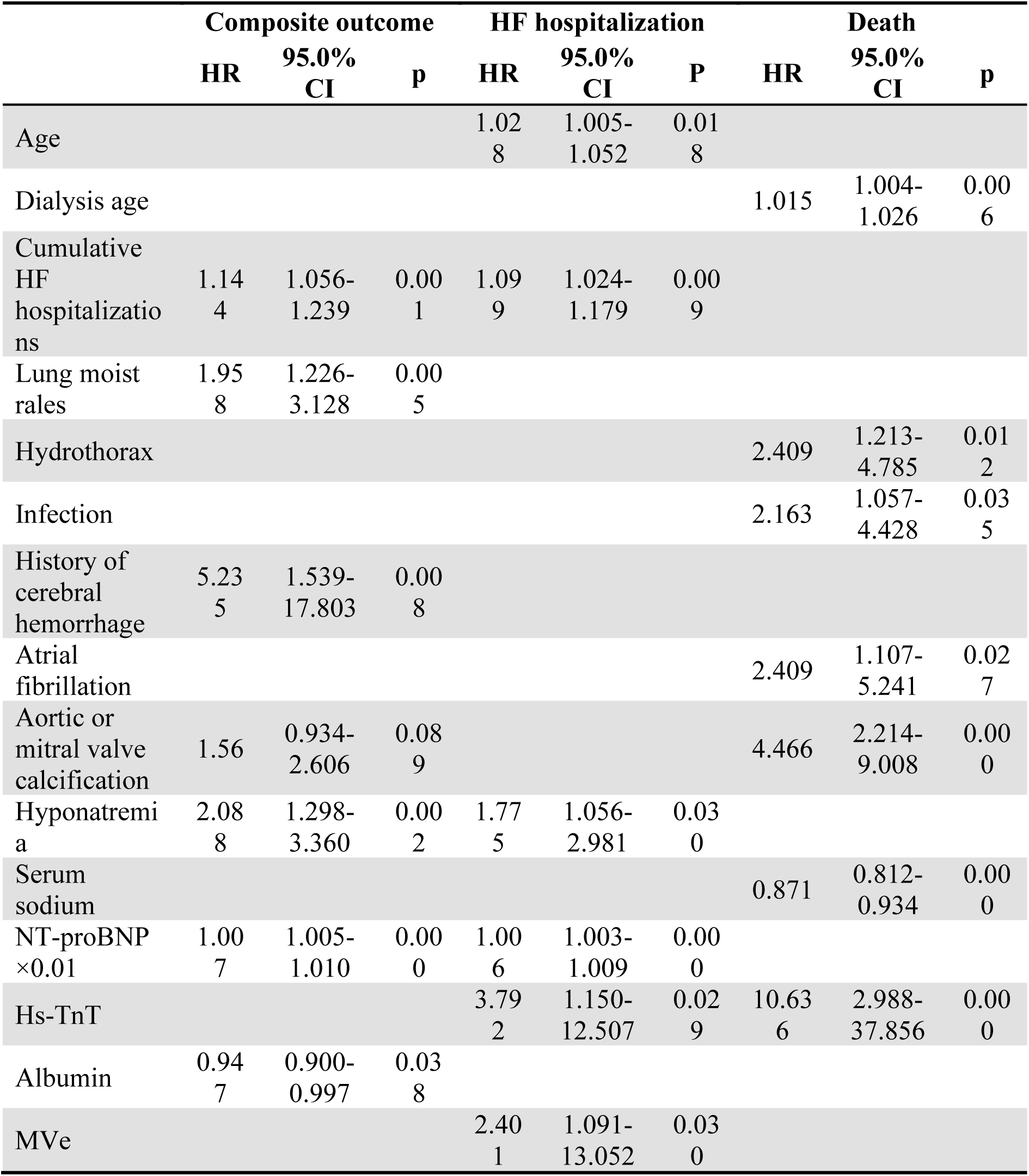
Multi-factor Cox model predictor distribution comparison.

**Figure S3:**
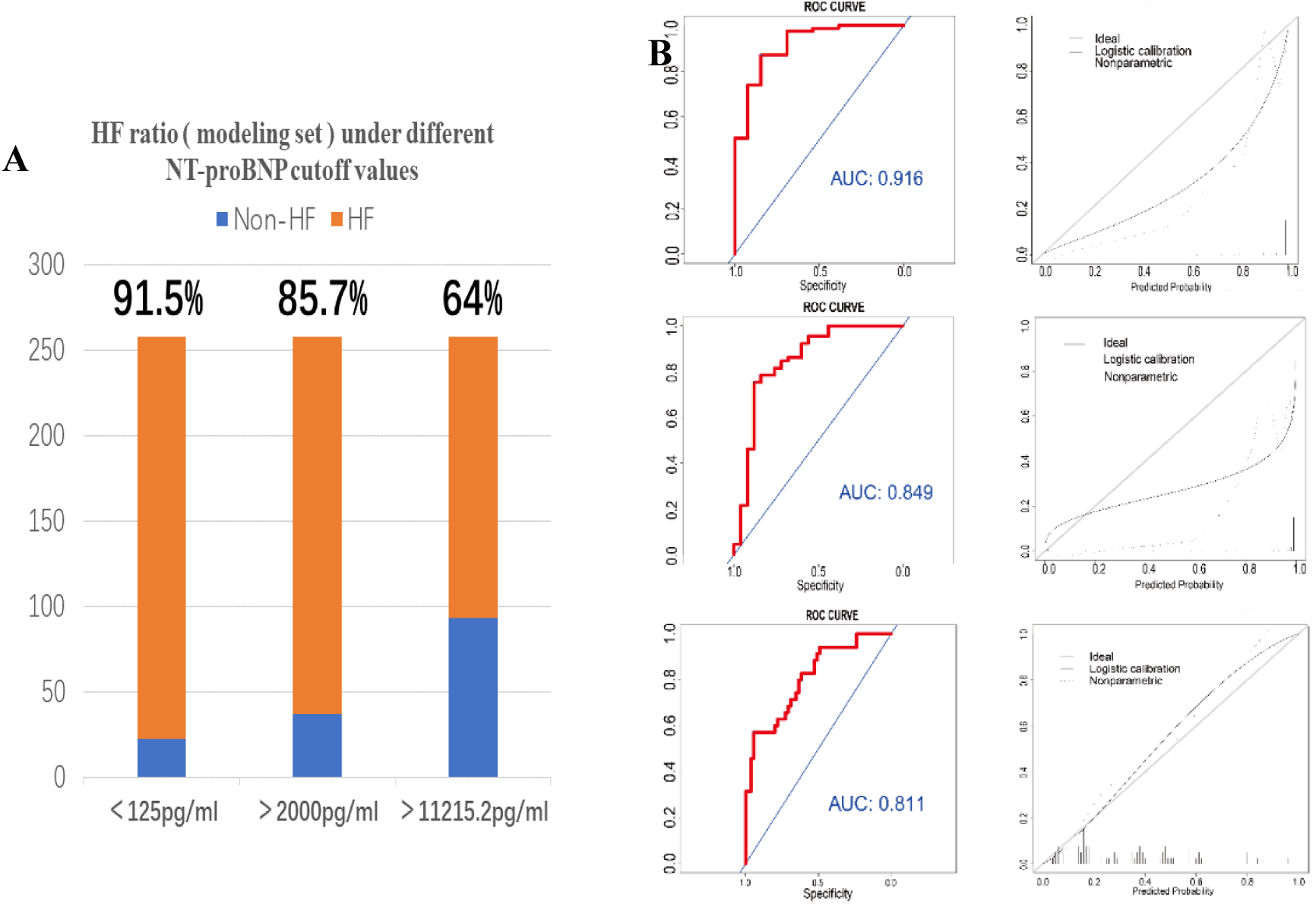
Diagnosis of heart failure and construction of logistic models according to different criteria: Diagnosis of heart failure and construction of logistic models according to different criteria: (A) The percentage of HF diagnosed with three different NT-proBNP cutoff values was significantly different. (B) Different NT-proBNP (from top to bottom: <125 pg/mL, > 2000 pg/mL, > 11 215.2 pg/mL) cut-off values were used to diagnose heart failure, construct a prediction model, and draw a calibration chart. It was suggested that the model constructed with NT-proBNP > 11215.2 pg/mL was clinically realistic.

**Table S4:**
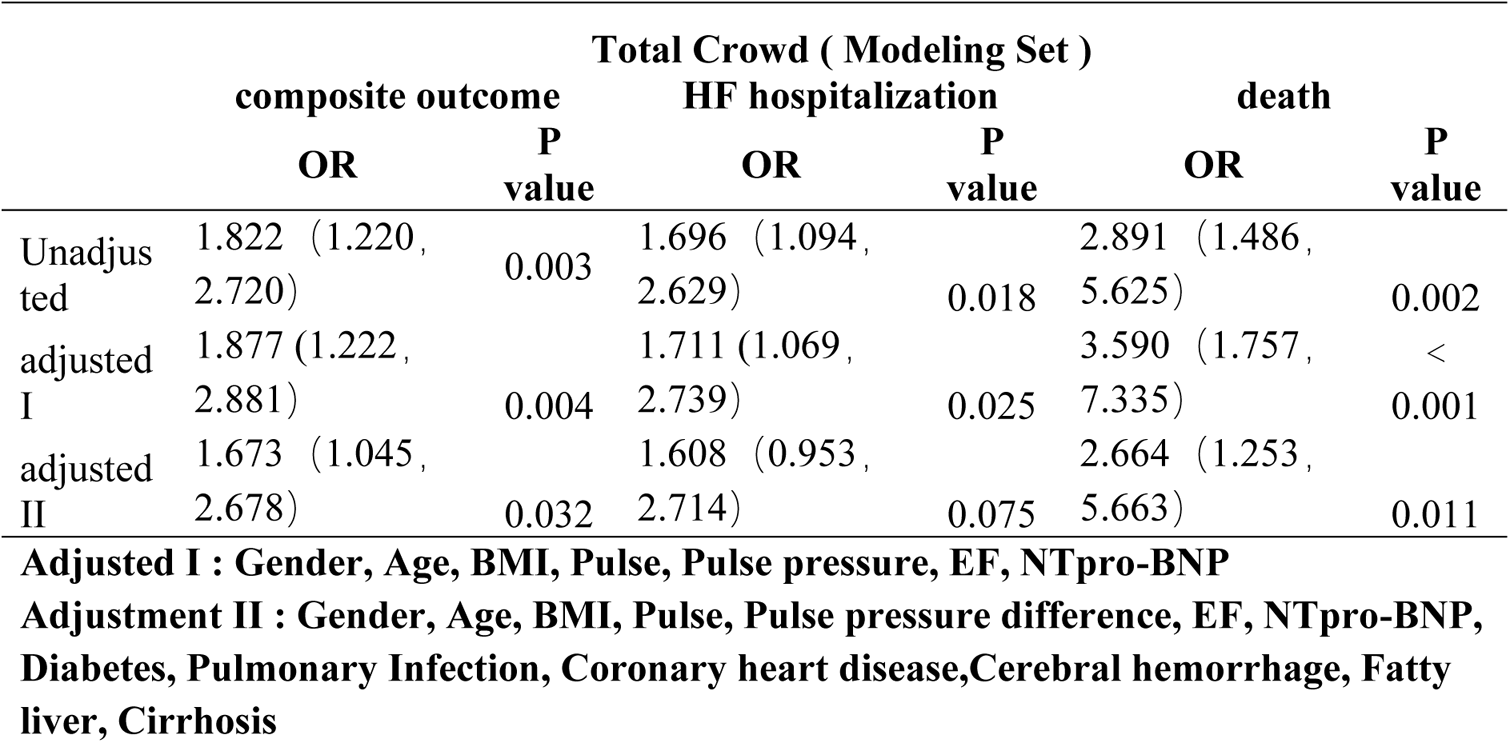
Association of hyponatremia with heart failure hospitalization and death after adjustment for comorbidities.

|  | Total Crowd ( Modeling Set ) |  |  |  |  |  |
| --- | --- | --- | --- | --- | --- | --- |
|  | composite outcome |  | HF hospitalization |  | death |  |
|  | OR | P value | OR | P value | OR | P value |
| Unadjusted | 1.822 (1.220, 2.720) | 0.003 | 1.696 (1.094, 2.629) | 0.018 | 2.891 (1.486, 5.625) | 0.002 |
| adjusted I | 1.877 (1.222, 2.881) | 0.004 | 1.711 (1.069, 2.739) | 0.025 | 3.590 (1.757, 7.335) | < 0.001 |
| adjusted II | 1.673 (1.045, 2.678) | 0.032 | 1.608 (0.953, 2.714) | 0.075 | 2.664 (1.253, 5.663) | 0.011 |
| **Adjusted I : Gender, Age, BMI, Pulse, Pulse pressure, EF, NTpro-BNP** |  |  |  |  |  |  |
| **Adjustment II : Gender, Age, BMI, Pulse, Pulse pressure difference, EF, NTpro-BNP, Diabetes, Pulmonary Infection, Coronary heart disease,Cerebral hemorrhage, Fatty liver, Cirrhosis** |  |  |  |  |  |  |

**Table S5:** A logistics model with pleural effusion as an outcome event and multiple comorbidities as predictors was constructed.

|  | Logistics single factor analysis |  |  | Logistics multi-factor analysis |  |  |
| --- | --- | --- | --- | --- | --- | --- |
|  | OR | 95%(CI) | p value | OR | 95%(CI) | p value |
| Coronary heart disease | 0.805 | (0.414-1.567) | 0.523 |  |  |  |
| Cerebral infarction or insufficient blood supply | 1.348 | (0.71-2.562) | 0.361 |  |  |  |
| Encephalorrhagia | 0.824 | (0.157-4.34) | 0.82 |  |  |  |
| Atrial fibrillation | 3.946 | (1.813-8.59) | 0.001 | 3.363 | (1.462-7.731) | 0.004 |
| Metabolic acidosis | 1.717 | (1.015-2.902) | 0.044 |  |  |  |
| Urinary tract infection | 0.679 | (0.179-2.5761) | 0.569 |  |  |  |
| Abnormal thyroid function | 0.549 | (0.149-2.022) | 0.367 |  |  |  |
| Diabetes | 1.015 | (0.601-1.715) | 0.955 |  |  |  |
| Diabetic Retinopathy | 0.944 | (0.502-1.775) | 0.858 |  |  |  |
| Lung infection | 2.95 | (1.723-5.051) | 0 | 2.365 | (1.315-4.256) | 0.004 |
| COPD | 1.587 | (0.737-3.416) | 0.238 |  |  |  |
| Respiratory failure<br>or pulmonary<br>edema | 6.84 | (1.801-25.98) | 0.005 | 5.477 | (1.364-21.997) | 0.017 |
| Bronchitis,<br>bronchiectasis | 1.334 | (0.578-3.081) | 0.499 |  |  |  |
| Emphysema or<br>pulmonary<br>aspergillosis | 1.457 | (0.7-3.032) | 0.314 |  |  |  |
| Silicosis,<br>pneumoconiosis,<br>lung destruction,<br>pulmonary<br>fibrosis | 4.3 | (0.772-23.965) | 0.096 |  |  |  |
| Fatty liver or<br>cirrhosis | 0.859 | (0.36-2.051) | 0.732 |  |  |  |
| Gallbladder stones<br>and post<br>cholecystectomy | 1.042 | (0.506-2.149) | 0.911 |  |  |  |
| Peptic ulcers,<br>inflammation of<br>the digestive tract | 0.913 | (0.5145-1.62) | 0.756 |  |  |  |
| Gastrointestinal<br>bleeding | 0.461 | (0.168-1.269) | 0.134 |  |  |  |
| Abnormal blood<br>clotting,<br>thrombocytopenia | 1.596 | (0.535-4.758) | 0.401 |  |  |  |
| Immunodeficiency | 0.512 | (0.056-4.654) | 0.552 |  |  |  |
| Tumors | 2.85 | (0.623-13.035) | 0.177 |  |  |  |
| Hyponatremia | 0.857 | (0.505-1.453) | 0.566 |  |  |  |
| HF | 5.915 | (2.936-11.915) | <<br>0.001 | 4.312 | (2.073-8.967) | < 0.001 |

